# Homologous recombination deficiency (HRD) can predict the therapeutic outcomes of immuno-neoadjuvant therapy in NSCLC patients

**DOI:** 10.1101/2022.03.12.22272306

**Authors:** Zhen Zhou, Zhengping Ding, Jie Yuan, Shengping Shen, Hong Jian, Qiang Tan, Yunhai Yang, Zhiwei Chen, Qingquan Luo, Xinghua Cheng, Yongfeng Yu, Xiaomin Niu, Liqiang Qian, Xiaoke Chen, Linping Gu, Ruijun Liu, Shenglin Ma, Jia Huang, Tianxiang Chen, Ziming Li, Wenxiang Ji, Liwei Song, Lan Shen, Long Jiang, Zicheng Yu, Chao Zhang, Zaixian Tai, Changxi Wang, Rongrong Chen, Shun Lu

**Author notes:** **Corresponding author:** Shun Lu, MD Shanghai Lung Cancer Center, Shanghai Chest Hospital, Shanghai Jiao Tong University, No. 241 West Huaihai Road, Xuhui District, Shanghai, 200030, China. These authors contributed equally to this work (Zhen Zhou, Zhengping Ding, Jie Yuan).

## Abstract

**Background:** Neoadjuvant immunotherapy is emerging as novel effective intervention in lung cancer but the study to prioritize effective surrogates indicating its therapeutic outcomes is limited. We investigated the genetic changes between patients with distinct response to neoadjuvant immunotherapy in non-small-cell lung cancer (NSCLC) for the derive of biomarkers with indicative capability in predicting outcomes.

**Methods:** In this study, 3 adenocarcinoma and 11 squamous cell carcinoma NSCLC patients were treated by neoadjuvant immunotherapy with variated regimen followed by surgical resection. Pre-therapy FFPE or fresh tissues and blood samples were analyzed by whole-exome sequencing (WES). Genetic alternation comparisons were conducted between differently-responded patients. Multiple public cohorts were selected for validation.

**Results:** DNA damage repair (DDR)-related InDel signatures and DDR-related gene mutations were enriched in better-responded patients, i.e. major pathological response (MPR) group. Besides, MPR patients exhibited provoked genome instability and unique homologous recombination deficiency (HRD) events. By further inspecting alternation status of homology-dependent recombination (HR) pathway genes, the clonal alternations were exclusively enriched in MPR group. Additionally, associations between HR gene alternations, percent of viable tumor cells and HRD event were identified, which orchestrated tumor mutational burden (TMB), mutational intratumor heterogeneity (ITH), somatic copy number alteration (SCNA) ITH and clonal neoantigen load in patients. Validations in public cohorts further supported the generality of our findings.

**Conclusions:** We innovatively associated the HRD event with enhanced neoadjuvant immunotherapy response in lung cancer. The power of HRD event in patient therapeutic stratification persisted in multi-facet public cohorts. We propose the inspection of HR pathway gene status could serve as novel and additional indicators directing immune-neoadjuvant and immunotherapy treatment decisions for NSCLC patients.

## Introduction

Being characterized by its high incidence and low curative rate, lung cancer remains the leading cause of cancer death worldwide. In the past decades, neoadjuvant therapy has provided extra treatment opportunities for lung cancer patients and substantially prolonged their survival^1^. As the understanding of programmed cell death protein 1 (PD-1)/programmed death-ligand 1 (PD-L1) regulatory axis deepened, neoadjuvant immunotherapy emerged to be a novel effective intervention in the treatment of resectable lung cancer. For example, in the phase III trial CheckMate-816 (NCT02998528) assessing efficacy of neoadjuvant nivolumab plus chemotherapy on surgically resectable early-stage (stage I–IIIA) NSCLC, the combination led to a MPR rate of 36.9% and pathological complete response (pCR) rate of 24%, significantly outperforms the chemotherapy alone arm.

However, only a few studies were reported to find effective surrogates indicating the therapeutic outcomes of immuno-neoadjuvant therapy in lung cancer, and to select best therapy responders. Genetic markers such as TMB, tumor neoantigen burden (TNB) or loss of heterozygosity (LOH) in human leukocyte antigen (HLA) regions were reported with some predictive power in indicating patients’ response to immunotherapy^2^. Nevertheless, it remains elusive that if these markers could also show predictive power in monitoring the therapeutic outcomes of lung cancer immuno-neoadjuvant therapy. Herein by performing whole-exome sequencing on 14 lung cancer patients receiving immuno-neoadjuvant therapy, we analyzed samples’ genetic changes and their association with patients’ response to treatment.

In result, multiple gene and cytoband level alternations were identified through comparing differently-responded patients. Higher levels of HRD were unprecedentedly discovered in patients with better response and lower percent of viable tumor cells after treatment, accompanied by clonal deactivation of HR pathway genes. Associations between HRD event and TMB, mutational as well as SCNA level ITH and clonal neoantigen load were also uncovered in immuno-neoadjuvant patients. Surprisingly, through analyses on gene alternations, TMB, TNB as well as patient survival using public cohorts, the association between HRD event and therapeutic efficacy persisted regardless of NSCLC histological subtypes or stages. These findings for the first time proposed the association between HRD and neoadjuvant immunotherapy outcomes in early-stage NSCLC and validated the power of HRD event in various NSCLC patient treatment guidance. Our results indicated the feasibility of HRD gene testing as independent and complementary indicators to the therapeutic outcomes in lung cancer immuno-neoadjuvant and immunological therapy, rendering it promising to guide treatment decisions and inform lung cancer disease management in an unparalleled perspective.

## Materials and methods

### Patient enrollment, specimen collection and sequencing data acquisition

A total of 3 adenocarcinoma and 11 squamous cell carcinoma NSCLC patients were accrued from 2018 to 2020 at Shanghai Chest Hospital with immuno-neoadjuvant therapy treatment and surgical resection. The demographics of recruited patients and treatment characteristics were listed in Table 1. Pre-therapy FFPE/fresh tissues were biopsied and peripheral blood mononuclear cell (PBMC) were isolated from blood samples for genomic DNA extraction by AllPrep DNA/RNA FFPE/Mini Kit (Qiagen) or CWE9600 Blood DNA kit (CWBiotech, China) and subsequent 100bp paired-end WES library construction. Final sequencing data was acquired by Geneplus-2000 sequencing platform (Geneplus, Beijing, China). Metrics including total read volume, Q30 percentage, GC content and duplication rate were used for quality assessment.

### Mutation calling, copy number alternation identification and signature analysis

After BWA (version 0.7.10) read alignment on hg19 reference genome and sample coverage filtration, MuTect (version 1.1.4) in GATK (version 4.0) pipeline identified somatic single nucleotide variants (SNVs), small insertions and deletions (InDels) by comparing tissue/PBMC specimens while GATK prioritized segment-level somatic copy number alterations (SCNA). Two rounds of filtration were conducted on SNVs and InDels, including reliability filtering by retaining variants with low frequency (<=0.01) in population, non-zero variant allele frequency (>=0.01), more supporting reads (>=3) and functional alternations (nonsense, in-frame/frame-shift small insertions and deletions, mutations in canonical splice-sites). Cancer associated genes^3^ were further extracted to conduct cancer-related filtration. As for the germline mutations, the SNPs and InDels were identified following workflows in GATK. Annotations including mutation types, resided transcripts and allele frequencies of the healthy population were conducted by Ensembl Varient Effector Predictor^4^. Functional mutations predicted as damaging and deleterious by PolyPhen^5^ and SIFT^6^ tools were initially retained. Later mutations with allele frequency>0.01 in all as well as the East Asian population from ExAC, 1000G and gnomAD databases were discarded in the further analyses. In addition, GISTIC (version 2.0) detected significantly altered regions in MPR and Non-MPR (MPR/Non-MPR) groups and an in-house algorithm^7^ employed tissue/PBMC read coverages’ statistical significance for the inference of focal level SCNA. As for mutational signatures, unfiltered somatic SNVs were fed into MutationalPatterns^8^ tool to derive their relative contribution to the base substitution spectrum and absolute contribution of COSMIC v3 SBS (single base substitution) mutational signatures. COSMIC v3 DBS (double base substitution) and ID (InDel) signature absolute exposures were quantified by Sigminer^9^. Quantification of existing seven SCNA signatures in curated samples was obtained by CNsignatures^10^.

### Intratumor heterogeneity measurement and SNV/SCNA clonality annotation

The genomic diversification denoted by intratumor heterogeneity (ITH) were measured at both somatic SNV and SCNA scope. The mutant-allele tumor heterogeneity (MATH) score^11^ was calculated using sample-wise variant allele frequency (VAF) on filtered mutations. SNV and SCNA data were jointly considered to estimate the cancer cell fraction (CCF), cancer ploidy, tumor purity and rescaled copy ratio by ABSOLUTE^12^ followed by model selection manual review. SNV was further annotated as clonal if its CCF upper 95% confidence interval>=1 and the clonal mutation probability was higher. Between-allelic rescaled copy number ratio comparison was conducted to exclude copy neutral LOH (CNLOH) segments and annotate SCNA segments using allelic subclonal information from ABSOLUTE outputs.

### HRD event quantification and HR pathway gene analysis

A NGS-based R package scarHRD^13^ was used to calculate three HRD metrics including telomeric allelic imbalance (TAI), loss off heterozygosity (LOH) and number of large-scale transitions (LST). Genes from HR pathway and core pathway^14^ were collected for the intersection with clonality-annotated SNV/germline mutations/SCNA.

### Neoantigen identification and HLA-LOH event evaluation

pVACseq^15^ was exploited for neoantigen prediction and the TNB was normalized as per-megabase neoantigen that passed filtration. The HLA-LOH event occurrence was determined by LOHHLA tool^7^ and the alleles with allelic imbalance p-value<0.05 was retained.

### Public data curation for validation

For multi-cohort data validation on the discriminative power of HRD in clinical treatments, data were retrieved from multiple immunotherapy and chemotherapy lung cancer datasets. More specifically, based on treatment scheme, the curated datasets were classified into neoadjuvant immunotherapy, immunotherapy, chemotherapy and multi-therapy categories. As for the neoadjuvant immunotherapy group, results from two studies utilizing WES in genomic characteristic profiling on NSCLC patients were collected^16, 17^ (denoted as N Engl J Med. 2018 and J Immunother Cancer. 2020). As for immunotherapy group, three WES datasets^18–20^ denoted as Cancer Cell. 2018, Science. 2015, Nat Genet. 2018 and one targeted sequencing dataset^21^ namely J Clin Oncol. 2018 were included. One small-cell lung cancer (SCLC) dataset^22^ named as Ann Oncol. 2016 was also included to validate the therapeutic prediction power of HRD in chemotherapy-treated SCLC. As a category in which datasets contain patients received distinct therapy types, targeted sequencing results on tissue^23^ (named as Cancer Discov. 2017) and blood^24^ samples (named as Nat. Med. 2018) were collected for treatment level comparisons in multiple therapy categories. Based on the sequencing result availability, alternation frequencies of HR pathway genes (i.e. HRD event) were calculated using functional mutations, mutation and neoantigen burdens were compared and survival differences were measured. Additionally, multiracial datasets^25, 26^ named as Sci. Rep. 2015, J Thorac Oncol. 2020, TCGA-LUAD (The Cancer Genome Atlas, lung adenocarcinoma) and TCGA-LUSC (lung squamous cell carcinoma) were collected either from papers or TCGA repository in cBioPortal database^27^ to investigate the alternation frequency of HR pathway genes in treatment-free multiethnic patients. Two datasets^14, 28^ containing pan-cancer HRD event analysis on TCGA data were also integrated for HRD frequency statistics.

### Statistical analysis

Both the two-sided and one-side Wilcoxon rank-sum tests were used for evaluating the group-level difference of continuous values including mutation relative contribution, SCNA, HRD levels, TMB and TNB between MPR and Non-MPR patients. When the group-wise comparisons were conducted on categorical data, Fisher’s exact test was applied. Correlations between genetic metrics and percentage of viable tumor cells were measured by Spearman’s correlation coefficient. As for survival analysis, the Kaplan-Meier (K-M) survival curves were generated by survminer package^29^ which applied log-rank test for survival time comparisons.

## Results

### Minor difference was observed in clinicopathological characteristics between patients with distinct response to therapy

The clinicopathologic characteristics of the patients enrolled were shown in Table 1 and patients were further categorized as MPR when the viable tumor cells in the resection specimen was fewer than 10%^30^, among which three patients achieved pCR. The median age of MPR/Non-MPR group was 62 (range, 55-68) and 60 (range, 44-68) accordingly with no sex differences. Most of the MPR patients were diagnosed with stage IIIA NSCLC while stage IIIA/IIIB patients were evenly distributed in Non-MPR group. All patients received immuno-neoadjuvant therapy but with varied regimen combinations (Suppl. Fig. 1A-B). Defined as the time interval between first treatment and surgery, neoadjuvant duration did not exhibit significant difference between patients with different response (Suppl. Fig. 1C). The median duration time was 83 (69-97) and 71 (50-92) days respectively. No correlation was observed between PD-L1 expression and percent of viable tumor cells (Suppl. Fig. 1D).

### Mutations in homologous recombination-related genes preponderated in MPR group

A total of 11598 SNVs and InDels were identified by WES, while 4243 and 1290 mutations retained through filtrations (Suppl. Fig. 1E). No obvious number difference was observed between MPR/Non-MPR and FFPE/Frozen tissues (Suppl. Fig. 1F). Mutation signature analysis was conducted on unfiltered mutations to decipher the possible etiological reasons behind distinct therapeutic response. As shown in Fig. 1A, the substitution spectrum plot displayed insignificant statistically difference between groups. By mapping mutations to known mutational signatures in COSMIC database (accessed in March 2021), SBS 4 associated with tobacco-smoking-induced cancer contributed to high proportions of mutations in two groups (Fig. 1B), consistent with their smoking history. Sorted by log2 contribution fold change between MPR/Non-MPR groups, InDel signatures ID6 and ID8 preponderated in MPR patients, both were attributed to DNA damage repair (DDR) mechanisms (Figure 1B). Regarding the relatively low absolute exposure value to known features, no obvious enrichment was observed on DBS signatures. Besides, a total of 293 mutations in 209 cancer driver genes were detected after filtrations, with *TP53* (13/14, 92.9%), *MUC16* (6/14, 42.9%), *CSMD3* (5/14, 35.7%) and *MUC4* (5/14,35.7%) most frequently mutated (Fig. 1C). No EGFR/ALK/LKB1 mutations were found in all patients. By comparing these genes with other public immunotherapy next-generation sequencing cohorts, *ZFHX3* and *PIK3CA* recurrently exhibited significantly higher mutation frequency in our MPR patients (Suppl. Fig. 1G). Contrarily, more of our Non-MPR patients possessed *NUP93* mutations (Suppl. Fig. 1G). Of particular interest, mutated tumor suppressor genes were enriched in diverse pathways (Fig. 1D), where genes from MPR patients were enriched in DDR and homologous recombination-related pathways like R-HSA-5693567 and R-HSA-6796648 (Suppl. Fig. 1H), insinuating the occurrence of HRD events in better-responded patients.

**Figure 1.**
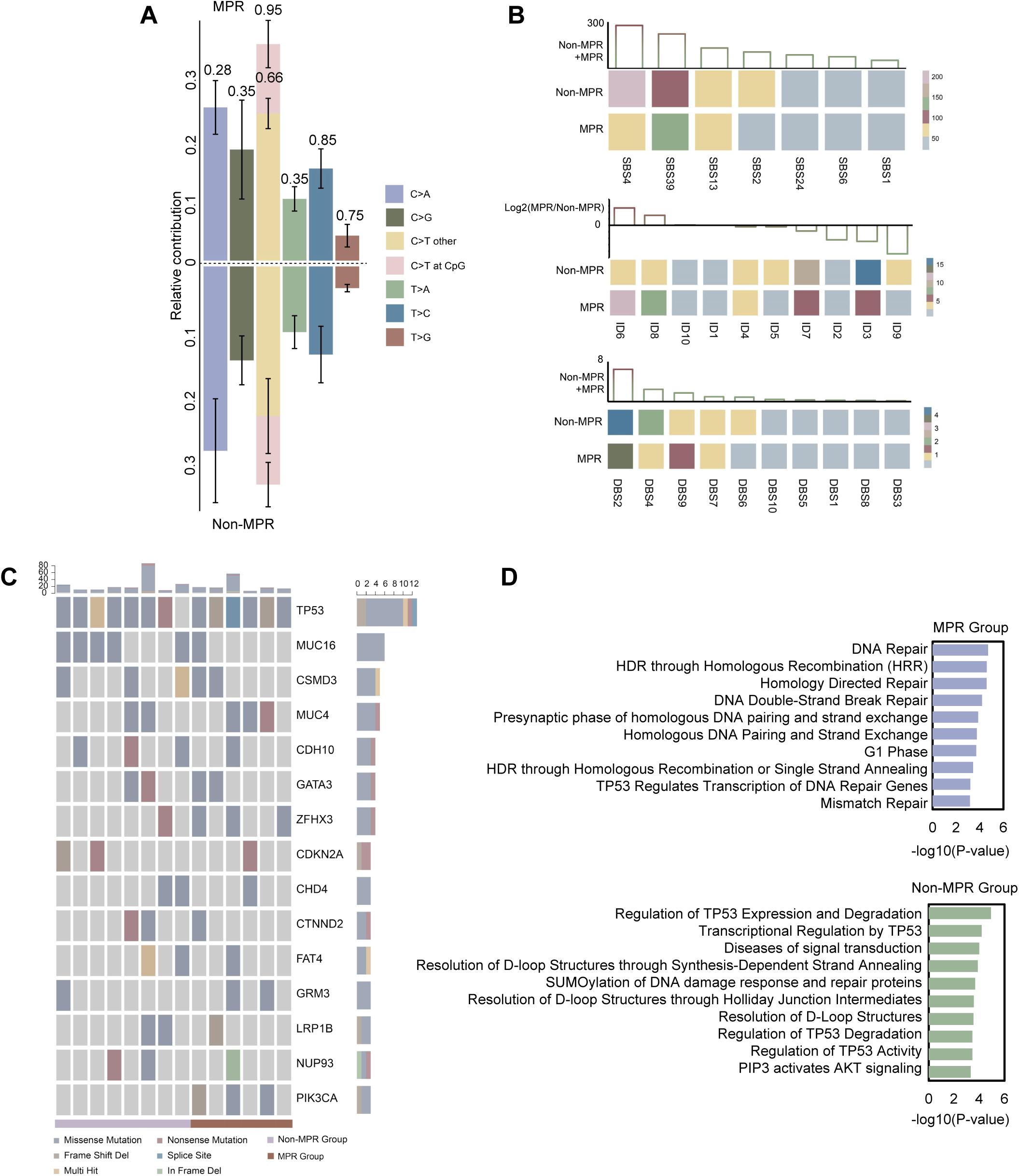
Mutational analysis results between MPR and Non-MPR groups. (A) Six substitution type spectrum plot in two patient groups. P-values were shown above each substitution category. (B) Exposures of three types of mutation signature to known database. SBS: single base substitution; ID: InDel; DBS: double base substitution. Either sum or log2 fold change of the absolute exposures was addedly depicted. (C) Landscape of frequent SNV and InDel on cancer driver genes. (D) Pathway enrichment analysis on mutated tumor suppressor genes.

### MPR group exhibited exacerbated somatic copy number alternations

Being the key mechanism repairing DNA double-strand breaks and interstrand crosslinks, alternations in HR pathway genes provoke genomic instability. Consistently, chromosome and arm level copy number variation (CNV) burden^31^ escalated in MPR group, both for all 14 patients (Fig. 2A-B) and squamous cell carcinoma (SCC) subgroup (Suppl. Fig. 2A-B). The two CNV burdens showed conspicuous negative correlation with percent of viable tumor cells (Fig. 2C-D). Percent of amplified or deleted genome also manifested an increased trend for all 14 patients (Suppl. Fig. 2C-E) and SCC subpopulation (Suppl. Fig. 2F-G). Among these altered segments in non-aneuploid samples, three SCNA signatures defined by CNsignatures tool exhibited different enrichment in MPR and Non-MPR group (Suppl. Fig. 2H). According to previous results from ovarian carcinoma^10^, the SCNA Signature 1 was anti-correlated with HRD SNV signature while SCNA Signature 7 showed positive association with HRD SNV signature 3. Unsurprisingly, the group-wise SCNA signature distributions showed high consistency with the mutation status of HR-related genes in MPR/Non-MPR patients (Suppl. Fig. 2I-J), i.e. lower Signature 1 and higher Signature 7 proportion in MPR group. Additionally, SCNA Signature 5, which was associated with subclonal copy number changes, got significantly elevated proportion in MPR group, raising the presumption that HR gene deactivation escalated SCNA level intratumor heterogeneity (ITH) in MPR group. When narrowing to SCC patients, the SCNA signature disparities reduced (Suppl. Fig. 2L-N). Considering copy number variation could be originated from multiple mechanisms including non-allelic homologous recombination (NAHR), non-homologous end joining (NHEJ), DNA replication error and meiotic processes^32^, SCNA signature analysis confirmed the association between HR pathway gene alternations and SCNA in our MIP subpopulation.

**Figure 2.**
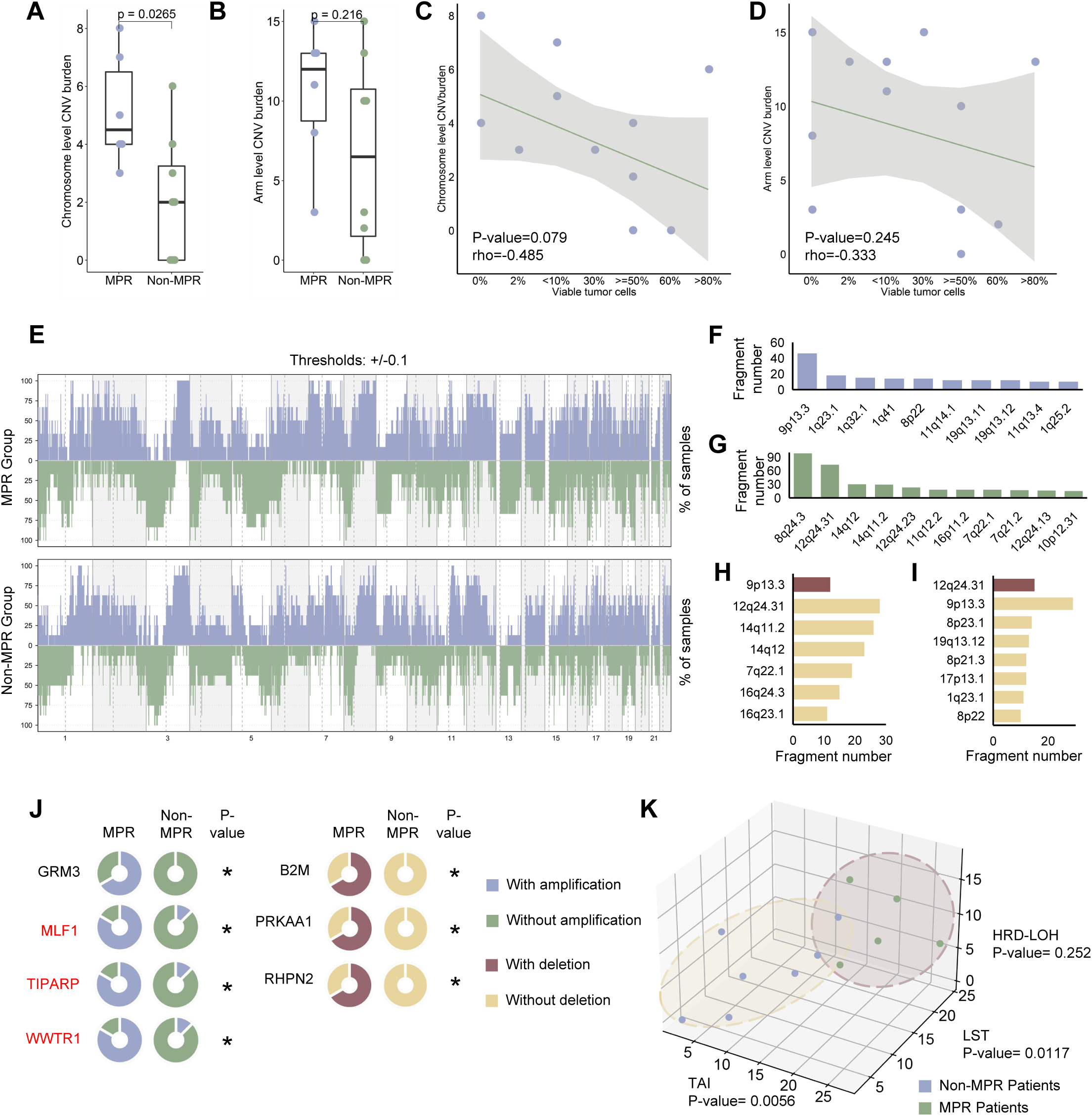
Analyses on copy number variations identified cytoband and gene-level SCNA difference in samples. (A) Chromosome level CNV burden between MPR/Non-MPR groups. (B) Arm level CNV burden in two groups. (C) Correlation between chromosome level CNV burden and percentage of viable tumor cells. (D) Correlation between arm level CNV burden and percentage of viable tumor cells. (E) Frequency plot of SCNA in two groups using copy number ratio threshold +/-0.1. (F) Top cytobands with significantly lower copy number ratio in MPR group. (G) Top cytobands with significantly higher copy number ratio in MPR group. (H) Top cytobands with higher sample-level amplification frequency differences in two groups. Alternations in Non-MPR group were marked with brown color. (I) Top cytobands with higher sample-level deletion frequency differences in two groups. Alternations in Non-MPR group were marked with brown color. (J) 7 genes were identified with significant focal SCNA occurrence difference between MPR/Non-MPR groups. Statistical significance was determined by Fisher’s exact test. *p<=0.03. Three genes with higher tumor to PBMC copy number difference were highlighted in red. (K) Three HRD metrics including TAI, LST and HRD-LOH calculated on non-aneuploid samples.

By testing the group-wise tumor to normal copy number ratio difference with Wilcoxon rank-sum test and sample-level copy number alternation frequency variance with Fisher’s exact test on genome-wide SCNA profile (Fig. 2E), we further pinpointed the recurrent SCNAs at cytoband level. Cytobands including 9p13.3, 1q23.1 and 1q32.1 possessed highest number of fragments with significantly lower copy number ratio in MPR group (Fig. 2F) while 8q24.3, 12q24.31 and 14q12 got most fragments with higher copy number ratio in MPR patients (Fig. 2G). Additionally, several cytobands were found with different sample-level copy number alternation frequency in two groups, among which cytobands 12q24.31 and 14q11.2 got highest amplification frequency in MPR group while 9p13.3 was most frequently amplified in Non-MPR samples (Fig. 2H). Opposingly, cytobands 9p13.3 and 8p23.1 tend to be frequently deleted in MPR patients while the deletion of 12q24.31 exhibited Non-MPR group-specificity (Fig. 2I). Interestingly, chromosome 9p13.3 was recurrently amplified in Non-MPR samples and deleted in MPR patients while chromosome 12q24.31 was recurrently deleted in Non-MPR group but amplified in MPR. We speculate the aberrant copy number of these cytobands could also provide hints in guiding NSCLC treatment.

To further discover the recurrent SCNAs at focal level, read coverages of tissue and PBMC in the two groups were statistically compared to identify genes altered in copy number with high confidence. 1589 genes were identified to harbor focal level SCNA, among which 7 genes including *GRM3, MLF1, TIPARP, WWTR1, B2M, PRKAA1* and *RHPN2* exhibited significantly different occurrence of focal alternations in the two groups, i.e. Fisher’s exact test P-value<=0.03 (Fig. 2J). When further filtered by higher tumor to PBMC copy number difference, genes *MLF1*, *TIPARP* and *WWTR1* were predominantly amplified in MPR group.

We also managed to highlight the significantly altered intra-group cytobands by GISTIC tool. As shown in Supplementary Figure 3A, multiple cytobands showed significant intra-group alternations. By further selecting group-specific cytobands (i.e. altered cytobands occurring only in MPR or Non-MPR patients), 24 cytobands were identified with group specificity (Suppl. Fig. 3A), containing 186 genes. 10 genes simultaneously harbored focal level SCNA (Suppl. Fig. 3B), among which 6 genes including *RAF1*, *CASC5*, *NTRK3, RAD51*, *SLC34A2* and *ZNF331* showed high consistency between cytoband level group specificity and focal level copy number alternation prevalence (Suppl. Fig. 3C).

Previous mutational analysis pinpointed the enrichment of alterations in HR genes in MPR group while SCNA signature quantifications associated the relationship between these alternations and SCNA. We next quantified the HRD-related event in non-aneuploid samples to confirm and compare its existence. Distributions of the three HRD-related events including telomeric allelic imbalance (TAI), large-scale state transitions (LST) and loss of heterozygosity (LOH) were significantly different between MPR/Non-MPR group (Fig. 2K). Unsurprisingly, when integrating the three HRD metrics, HRDscore in MPR group was significantly higher and negatively correlated with percent of viable tumor cells, both on all non-aneuploid samples (Suppl. Fig. 3D-E) and SCC samples (Suppl. Fig. 3F-G).

### Intratumor heterogeneity divergence caused by HRD potentially engendered the therapeutic response difference between two groups

Emergent evidence suggests that ITH impacts the effectiveness of immunotherapies^33^. Indeed, lower mutant-allele tumor heterogeneity (MATH) scores (i.e. lower ITH) were observed in MPR group (Suppl. Fig. 4A), which positively correlated with percentage of viable tumor cells (Suppl. Fig. 4B). Both the trends persisted in SCC samples (Suppl. Fig. 4C-D). Unsurprisingly, non-aneuploid and high purity MPR samples carried higher TMB values (Fig. 3A) and the TMB values were negatively correlated with percentage of viable tumor cells (Fig. 3B). Similar conclusions could be drawn when restricted to SCC samples (Suppl. Fig. 4E-F). When considering the clonality of mutations, disparity between MPR/Non-MPR group increased to significant (Fig. 3C) and clonal TMB value decreased with worse therapeutic response (Fig. 3D). Again, similar phenomena were observed on SCC subpopulation (Suppl. Fig. 4G-H). MPR patients also possessed relatively higher clonal SNV percentage (Suppl. Fig. 4I) and the better-responded ones got lower mutational inner heterogeneity (Suppl. Fig. 4J). Intriguingly, apart from the lower ITH, clonal somatic mutations in HR pathway genes were exclusively enriched in MPR group regardless of NSCLC histological subtype (Fig. 3E, Fisher’s exact test) and the only MPR patient without HR gene mutations had clonal base alternation in *ATR* gene, which essentially serves as a key player in DNA damage response and orchestrates the upstream signaling of HR^34^. In contrast, germline HR gene mutations did not exhibit an inconsonant trend (Suppl. Fig. 4K). The reduced ITH and enrichment of HR pathway gene mutations in MPR group suggested the possible inner association between HRD and mutational ITH, the impact of HRD events in pharmacodynamics of immuno-neoadjuvant therapy and the possibility of applying HRD events as markers guiding immunological treatment decisions for NSCLC patients.

**Figure 3.**
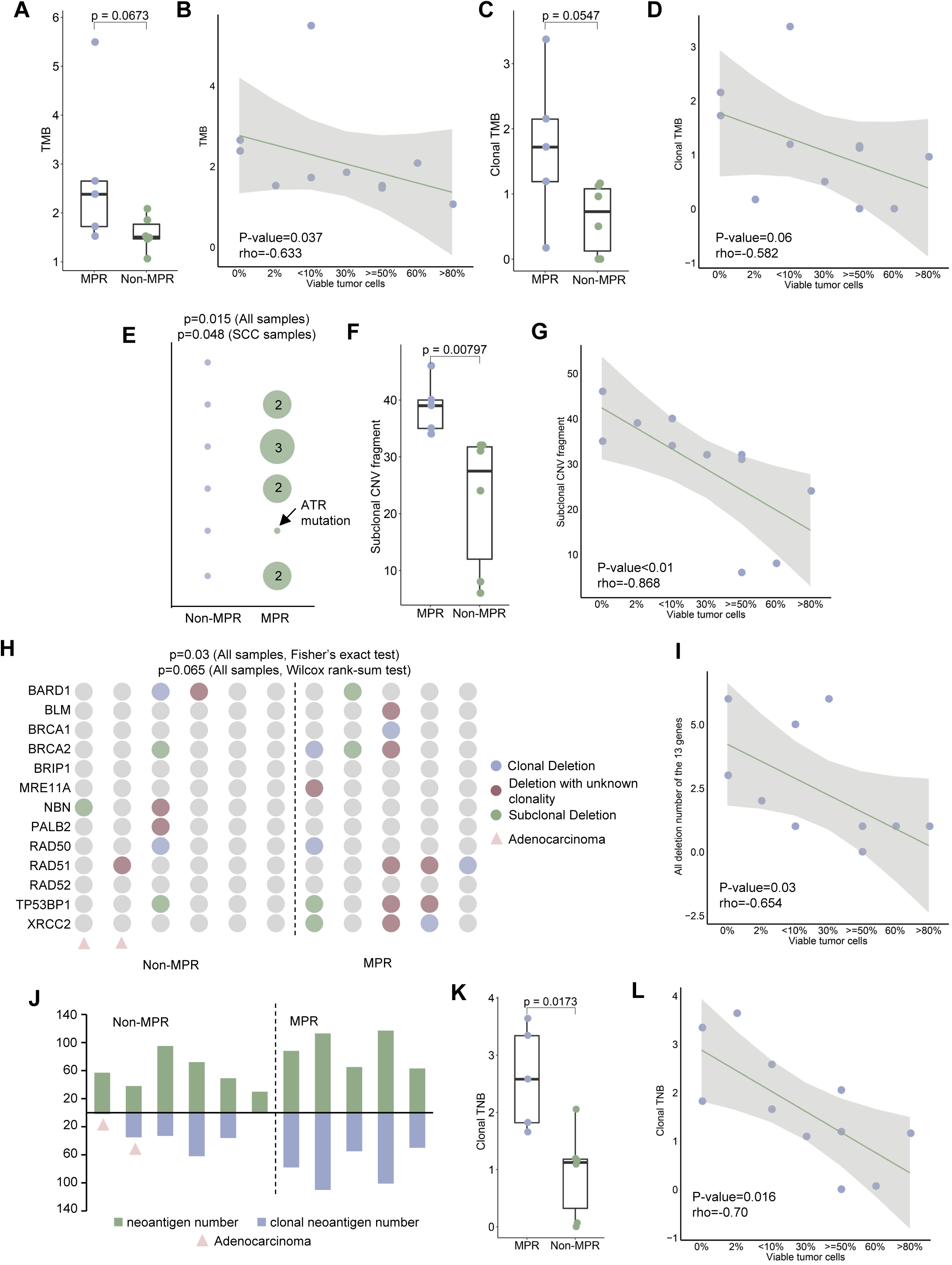
Clonal HR pathway gene deactivation impacts neoadjuvant immunotherapy consequences in NSCLC patients. (A) TMB value distribution between MPR/Non-MPR groups. (B) Correlation between TMB and percentage of viable tumor cells. (C) Clonal TMB value in two groups. (D) Correlation between clonal TMB and percentage of viable tumor cells. (E) Existence of HR pathway genes’ clonal SNV in two groups. The size of the circle was proportional to the mutation number. P-values were calculated by Fisher’s exact test on all and SCC samples. (F) Subclonal SCNA fragment number in two groups. (G) Correlation between subclonal SCNA fragment number and percentage of viable tumor cells. (H) Deletion status of 13 HR core pathway genes in MPR/Non-MPR groups. P-values were calculated by Fisher’s exact test or one-side Wilcoxon rank-sum test. Adenocarcinoma samples were marked with pink triangles. (I) Correlation between focal deletion number of the 13 genes and percentage of viable tumor cells. (J) All and clonal neoantigen number generated by two groups. Adenocarcinoma samples were marked with pink triangles. (K) Clonal TNB value distribution in two groups. (L) Correlation between clonal TNB and percentage of viable tumor cells.

Previous analysis on SCNA signatures implicated the possible existence of subclonal SCNA in MPR group, we further confirmed the conjecture by the SCNA landscape of non-aneuploid and high purity samples annotated by clonality (Fig. 3F). With increased percentage of viable tumor cells in the sample, the subclonal SCNA fragment number manifested a decreased trend (Fig. 4G), regardless of the tumor subtype (Suppl. Fig. 4L-M). Given that HRD essentially leads to the occurrence of recurrent SCNAs^35^ and the strong association between HRD and SCNA in our sample, the clonal HR mutations preponderated in MPR group could possibly be the source of these SCNA level ITH. Apart from mutational silence, a total of 13 core HR pathway genes’ deletion was sporadic in Non-MPR but concentrated in MPR patients (Fig. 3H) with statistical significance (P-value=0.03, Fisher’s exact test). The deletion events also negatively correlated with the percentage of viable tumor cells (Fig. 3I). In addition, proportion of genes possessing clonal deletion was also higher in MPR (P-value=0.065, one-side Wilcoxon rank-sum test) and clonal deletion events vanished with increased proportion of viable tumor cells (Suppl. Fig. 4N), indicating activities of HR genes could be synergistically altered at mutation and CNV level, consequently resulting in the variance of immuno-neoadjuvant clinical outcome in NSCLC.

**Figure 4.**
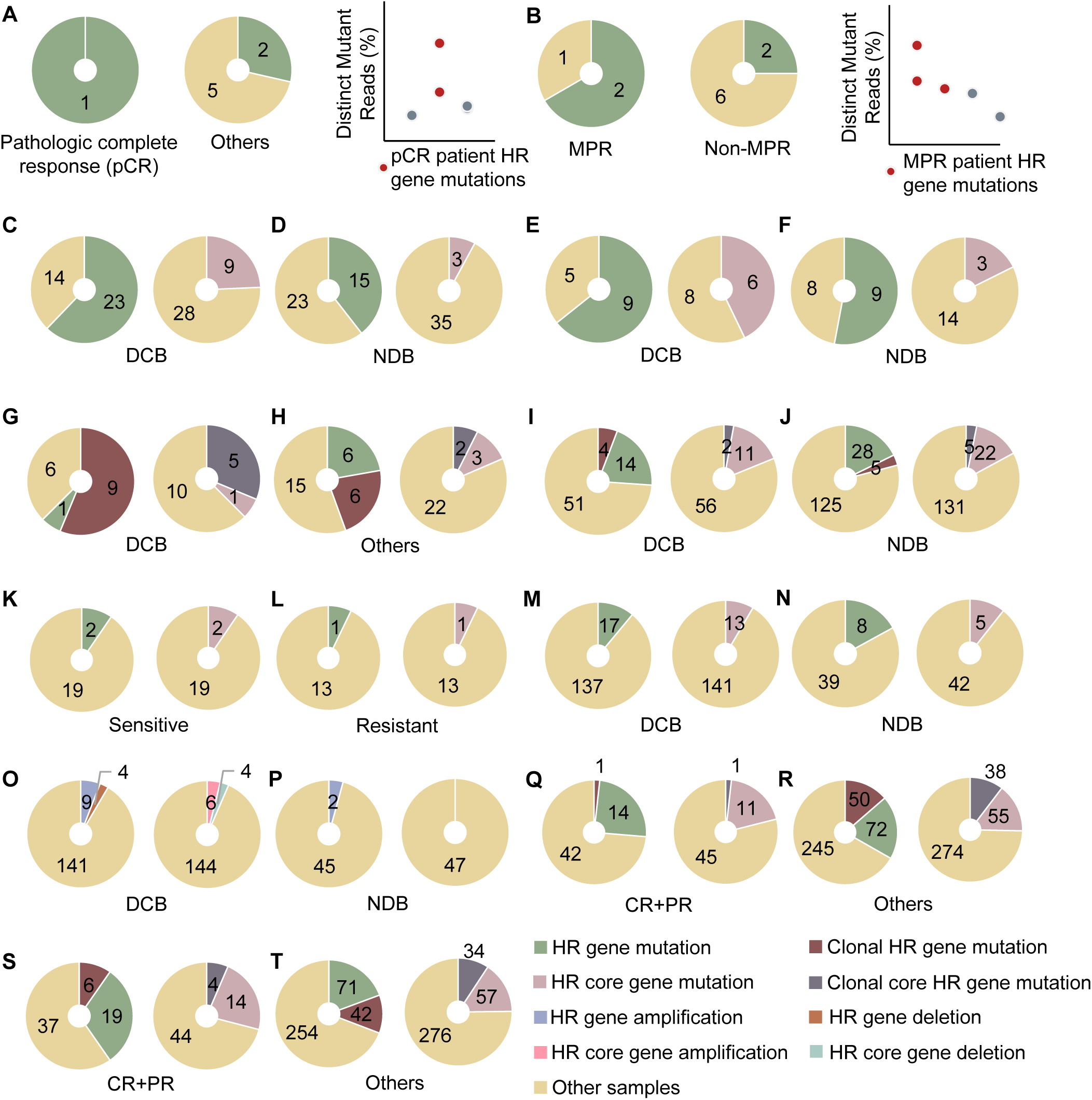
Investigation on HR pathway gene alternation frequency in patients with distinct response from public cohorts. (A) HR pathway gene mutation frequency in J Immunother Cancer. 2020 neoadjuvant dataset. Left: in pCR patients. Middle: in other patients. Right: percentage of distinct mutant reads for each HR gene mutation detected. Mutations from pCR patients were marked with red color. (B) HR pathway gene mutation frequency in N Engl J Med. 2018 neoadjuvant dataset. Left: in MPR patients. Middle: in Non-MPR patients. Right: percentage of distinct mutant reads for each HR gene mutation detected. Mutations from MPR patients were marked with red color. (C) HR pathway gene mutation frequency in DCB patients from Cancer Cell. 2018 dataset. Left: HR gene mutations. Right: HR core pathway mutations. (D) Similar with (C), but in NDB patients. (E) Similar with (C), but using Science. 2015 dataset. (F) Similar with (D), but using Science. 2015 dataset. (G) HR pathway gene mutation frequency in DCB patients from Nat Genet. 2018 dataset. Left: all and clonal HR gene mutations. Right: all and clonal HR core pathway mutations. (H) Similar with (G), but using non-DCB patients. (I) Similar with (G), but using J Clin Oncol. 2018 dataset. (J) Similar with (I), but using NDB patients. (K) HR pathway gene mutation frequency in chemo-sensitive patients from Ann Oncol. 2016 dataset. Left: HR gene mutations. Right: HR core pathway gene mutations. (L) Similar with (K), but in chemo-resistant patients. (M) HR pathway gene mutation frequency in patients achieved DCB in chemotherapy from Cancer Discov. 2017 dataset. (N) Similar with (M), but in NDB patients. (O) HR pathway gene SCNA frequency in patients achieved DCB in chemotherapy from Cancer Discov. 2017 dataset. Left: HR gene SCNAs. Right: HR core pathway gene SCNAs. (P) Similar with (O), but in NDB patients. (Q) HR pathway gene mutation frequency in patients achieved CR and PR in chemotherapy from Nat Med. 2018 dataset. Left: all and clonal HR gene mutations. Right: all and clonal HR core pathway mutations. (R) Similar with (Q), but in other patients. (S) Similar with (Q), but using patients received immunotherapy. (T) Similar with (S), but in other patients.

We next scrutinized the number of neoantigens generated in two groups. As expected, after discarding aneuploid or low purity samples, MPR group generated higher amount of neoantigens in all patients and SCC subgroup with higher clonal proportion (Fig. 3J). Similarly, TNB values in MPR group was higher for all patients (Suppl. Fig. 5A) and negatively correlated with percentage of viable tumor cells (Suppl. Fig. 5B). These trends persisted in SCC subpopulation (Suppl. Fig. 5C-D). Unsurprisingly, the clonal TNB value measuring neoantigen-level ITH was significantly higher in MPR subgroup when considering all patients (Fig. 3K) or only SCC ones (Suppl. Fig. 5E), which possibly brought by low mutational level ITH. Expectedly, neoantigens load was inversely correlated with percentage of viable tumor cells in sample, both for all patients (Fig. 3L) and SCC subtype (Suppl. Fig. 5F).

Besides the total amount, neoantigen load could be concomitantly affected by the activity of HLA class I genes. Unsurprisingly, the LOH frequency of HLA-A, HLA-B and HLA-C loci in Non-MPR group was higher (Suppl. Fig. 5G). When only considering neoantigens on the kept HLA alleles, MPR patients owned higher neoantigen load (Suppl. Fig. 5H). Again, the amount of the kept neoantigens decreased with percentage of viable tumor cells (Suppl. Fig. 5I) and such tendency persevered in SCC patients (Suppl. Fig. 5J-K). As for low-ITH neoantigens on kept HLA alleles, differences between MPR and Non-MPR group exacerbated in all patients (Suppl. Fig. 5L) or SCC population (Suppl. Fig. 5N). Analogous correlation trend between kept clonal TNB and quantified therapeutic response was also observed (Suppl. Fig. 5M, O). Astoundingly, these HLA-retained clonal neoantigens were mostly resided in the regions with somatic copy number alternations in MPR group (Suppl. Fig. 6A) and these CNV-related neoantigen was associated with percent of variable tumor cells (Suppl. Fig. 6B). The 9 SCC patients also demonstrated a similar trend (Suppl. Fig. 6C-D). When taking the alternation type into account, the clonal neoantigens on kept HLA alleles were predominantly resided in the amplified segments (amplification type SCNA) for MPR patients (Suppl. Fig. 6E), denoting an inner association between clonal TNB and copy number amplification on our samples. The kept clonal neoantigen number on amplified segments demonstrated akin negative correlations with percentage of viable cells regardless of histological subtype (Suppl. Fig. 6F-G). Notably, the only MPR sample without HR gene mutations possessed the highest number of clonal neoantigen (n=110) but they scarcely resided in SCNA segments (Suppl. Fig. 6E), prompting the hypothesis that mutations in HR genes lead to the increased SCNA ITH and LOH of HLA, and eventually orchestrate to compensate the amount of clonal antigen burden in patients, ultimately enhancing the response to immuno-neoadjuvant therapy. In conclusion, these findings offer potential strategies for prioritizing NSCLC patients who could benefit the most from immuno-neoadjuvant therapy.

### Multi-cohort validation confirmed the feasibility of HRD mutational testing in immunotherapy patient stratification

Previous analyses confirmed the association between HRD event or HR pathway gene alternation and key determinants of neoadjuvant immunotherapy outcome in all 11 non-aneuploid and high purity samples (Suppl. Fig. 7A). Moreover, HRD event correlated well with clonal neoantigen generation and CNV-related clonal neoantigens in MPR samples (Suppl. Fig. 7B) and such correlations alleviated in Non-MPR patients (Suppl. Fig. 7C). To further confirm the generalized power of HR pathway gene alternation in therapeutic outcome prediction, two neoadjuvant immunotherapy cohorts, four immunotherapy cohorts, one chemotherapy cohort and two cohorts containing patients received distinct therapy types were curated for validation (details in Materials and Methods). As shown in Supplementary Table 1, the selected datasets variated in therapeutic regimens and treatment design. To begin with, we firstly scrutinized the alternation frequency of HR pathway genes in distinctly-responded patients from different public cohorts. For J Immunother Cancer. 2020 dataset from the two neoadjuvant studies, all patient (100%) achieved pCR had HR pathway gene mutation (Fig. 4A, left) while the HR gene mutation was sporadic in other patients (Fig. 4A, middle, 28.5% patients). When inspecting HR gene mutation clonality, pCR patients possessed mutations with higher percentage of distinct mutant reads (Fig. 4A, right), indicating the lower HR gene mutational ITH in better-responded subgroup. Similar high prevalence of HR gene mutations in MPR patients (Fig. 4B, left, 66.7% patients), the HR gene mutational disperse in Non-MPR group (Fig. 4B, middle, 25% patients) as well as lower HR gene-related ITH (Fig. 4B, right) were observed in N Engl J Med. 2018 neoadjuvant dataset. As for the immunotherapy datasets, patients with durable clinical benefit (DCB) possessed higher HR gene (Fig. 4C, E, left) or HR core pathway gene mutation frequency (Fig. 4C, E, right) when compared to no durable benefit (NDB) subpopulation (Fig. 4D, F) in both Cancer Cell. 2018 and Science. 2015 cohorts. After addedly considering mutational ITH by annotating mutations with somatic clonal probability>=0.8 as clonal, DCB patients from Nat Genet. 2018 cohort significantly carried more clonal HR gene mutations (P-value=0.045, Fisher’s exact test) and clonal HR core pathway gene mutations (P-value=0.08, Fisher’s exact test) than other patients (Fig. 4G-H). Interestingly, such discrepancy was not observed in targeted sequencing immunotherapy dataset J Clin Oncol. 2018 (Fig. 4I-J), denoting the prominence of panel design in targeted sequencing when using HR pathway genes’ mutational status as immunotherapy biomarkers. In addition, for metastatic SCLC patients from Ann Oncol. 2016 dataset, no obvious proportional difference on mutated HR genes was observed between chemo-sensitive and chemo-resistant groups (Fig. 4K-L), denoting the vague connection between HR gene mutation and chemotherapy response in metastatic SCLC. Similar association between HR pathway alternations and treatment could be derived in NSCLC patients with distinct response to chemotherapy in Cancer Discov. 2017 cohort (Fig. 4M-N) and focal SCNA frequency of HR pathway genes among these patients exhibited an analogous trend (Fig. 4O-P). Lastly, for the blood-based targeted sequencing dataset Nat Med. 2018, patients achieved complete response (CR) or partial response (PR) in chemotherapy and other chemo-treated patients showed similar HR gene mutation frequency (Fig. 4Q-R). When inspecting patients received immunotherapy, HR gene mutational prevalence increased slightly in CR+PR (CR and PR) patients (Fig. 4S-T). In conclusion, multi-cohort validation confirmed the increased HR pathway gene mutational frequency in patients better-responded to immunotherapy while HR genes demonstrated negligible connection with therapeutic outcomes in chemo-treated lung cancer patients. When applying HR pathway gene alternations as immunotherapy biomarkers in targeted sequencing, extra attention should be given to panel design.

Additionally, regarding the availability of data, comparisons on genomic characteristics were further conducted between patients stratified by therapeutic response and HR pathway gene conditions. More specifically, four immunotherapy datasets including Cancer Cell. 2018, Science. 2015, Nat Genet. 2018, J Clin Oncol. 2018 and Nat Med. 2018 cohort containing chemo/immuno-treated subgroups were selected for comparisons on TMB, TNB and patient survival. To begin with, TMB values in non-squamous DCB patients from Cancer Cell. 2018 dataset was significantly higher than NDB group (Fig. 5A, left) while TMB failed to distinguish squamous DCB and NDB patients (Fig. 5A, right). Similarly, TMB and clonal TMB (defined on mutations with tumor allele frequency (Taf)>=0.2) elevated in DCB subgroup in Science. 2015 dataset (Fig. 5B). Unsurprisingly, patients in Science. 2015 with HRD (HR pathway gene mutations) and clonal HR gene mutations possessed significantly higher clonal TMB value (Fig. 5C). In non-squamous patients from Nat Genet. 2018 dataset, we observed congruent TMB and clonal TMB distributions between groups stratified by curative effect (Fig. 5D) or HR gene clonal mutations (Fig. 5E), again endorsed the power of HRD in therapeutic effect prediction. As for DCB and NDB patients from J Clin Oncol. 2018 cohort, though TMB elevated in DCB subpopulation (Suppl. Fig. 8A), it again failed to distinguish squamous patients with different response (Fig. 5F, right). When inspecting patients with and without HR gene mutations, the TMB disparity increased for both adenocarcinoma (Fig. 5G, left) and squamous (Fig. 5G, right) patients. Moreover, patients with HR gene mutations in J Clin Oncol. 2018 cohort possessed higher clonal TMB (Fig. 5H, left) while HR gene clonal mutations (defined by Taf>=0.3) prioritized patients with significantly higher TMB (Suppl. Fig. 8B) and clonal TMB (Fig. 5H, right). Finally, for immunotherapy patients in the biomarker-evaluable population of Nat Med. 2018 dataset, CR+PR patients got significantly higher TMB value (Suppl. Fig. 8C) but TMB again failed to stratify the better-responded patients in squamous subpopulation (Fig. 5I, right). Intriguingly, CR+PR patients with HRD bore higher TMB than other CR+PR patients (Fig. 5J) and patients with HR gene global or clonal (defined by Taf>=0.05) mutations in blood unsurprisingly harbored higher clonal TMB (Fig. 5K). The above comprehensive analysis on TMB confirmed their substantial elevation in HRD patients receiving diversified non-neoadjuvant treatment regimens and possessing varying clinicopathologic characteristics. Regarding higher TMB correlates with better-responded NSCLC patients, our observations sustained the feasibility of using HRD or HR gene mutations as novel immunotherapy biomarker in NSCLC.

**Figure 5.**
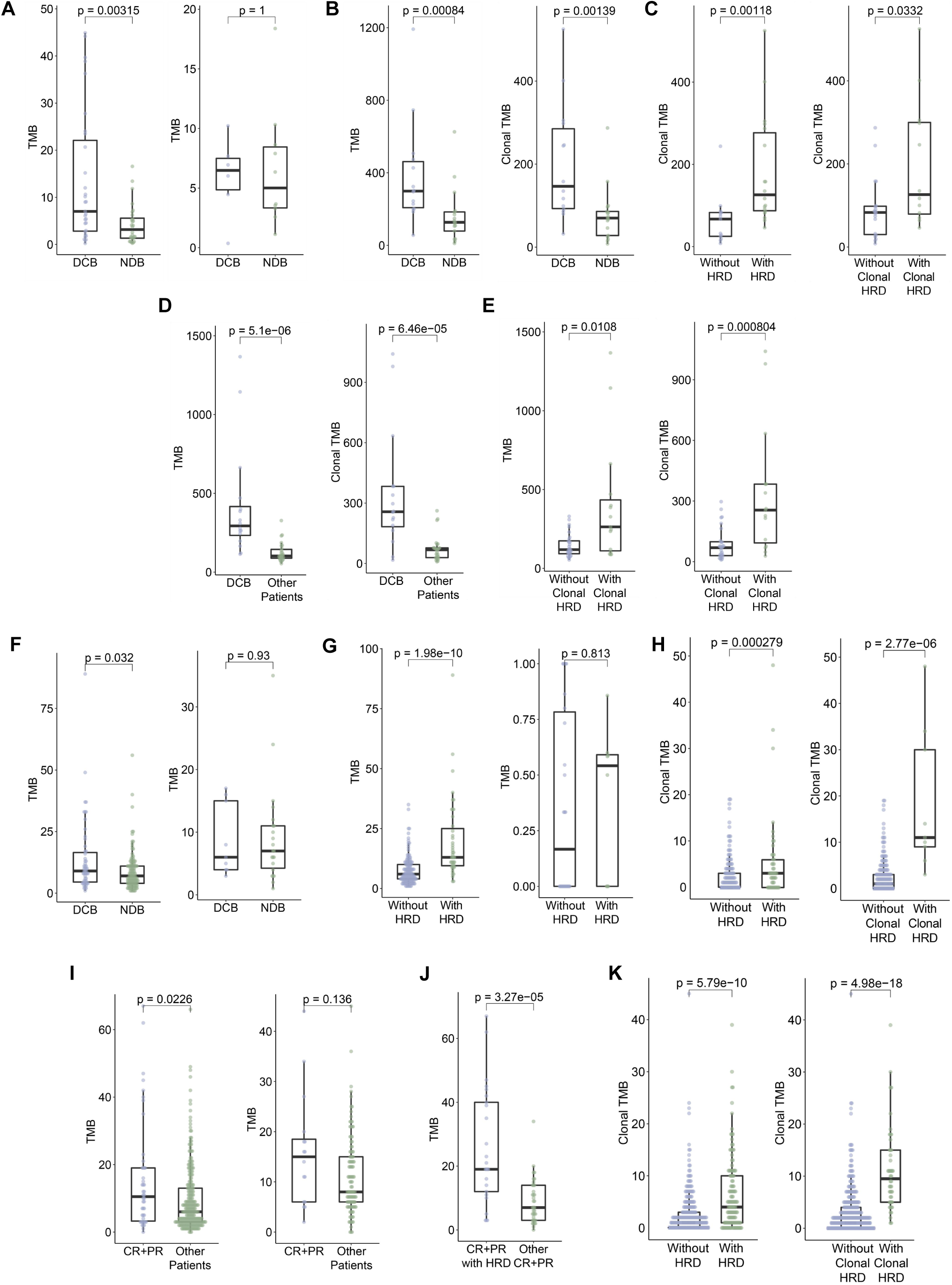
Comparisons on TMB values stratified by therapeutic response and HR pathway gene status from public cohorts. (A) TMB value distribution between DCB and NDB patients in Cancer Cell. 2018. Left: in non-squamous subgroup. Right: in squamous subgroup. (B) TMB and clonal TMB value distribution between DCB and NDB patients in Science. 2015. Left: TMB. Right: clonal TMB. (C) Clonal TMB value distribution between patients stratified by HRD event in Science. 2015 dataset. Left: between patients with and without HR pathway gene mutation. Right: between patients with and without HR pathway gene clonal mutation. (D) TMB and clonal TMB value distribution between DCB and other patients in non-squamous population from Nat Genet. 2018 dataset. Left: TMB. Right: clonal TMB. (E) Similar with (D), but using HR pathway gene clonal mutations as the stratification strategy. (F) TMB value distribution between DCB and NDB patients in J Clin Oncol. 2018. Left: in adenocarcinoma subgroup. Right: in squamous subgroup. (G) Similar with (F), but using HR pathway gene mutation as the classification strategy. (H) Similar with (C) but in J Clin Oncol. 2018 dataset. (I) TMB value distribution between CR+PR and other immunotherapy patients in Nat Med. 2018 dataset. Left: in non-squamous subgroup. Right: in squamous subgroup. (J) TMB value distribution between CR+PR patients with HRD and other CR+PR patients in Nat Med. 2018. (K) Similar with (H), but in Nat Med. 2018 dataset.

We next anatomized the TNB and clonal TNB distribution between patients assorted by treatment outcome or HR pathway gene mutational condition. Firstly, for Cancer Cell. 2018 dataset, TNB and clonal TNB was significantly higher in HRD (HR pathway gene-mutated) group (Suppl. Fig. 8D) and such trend preserved in non-squamous (Fig. 6A) and squamous (Fig. 6B) NSCLC histological subtypes. Likewise, apart from the significant elevation of TNB and clonal TNB in DCB group (Suppl. Fig. 8E), patients harboring HR pathway gene global and clonal mutations in Science. 2015 cohort generated more neoantigens (Fig. 6C-D). Concerning the aggrandize of neoantigen number as well as heterogeneity provided surety for immuno-therapeutic benefits, these results on non-neoadjuvant cohorts shared high consistency with conclusions in our patients and again advocated the use of HRD event which chaperoned TNB incline in immunotherapy patient stratification.

**Figure 6.**
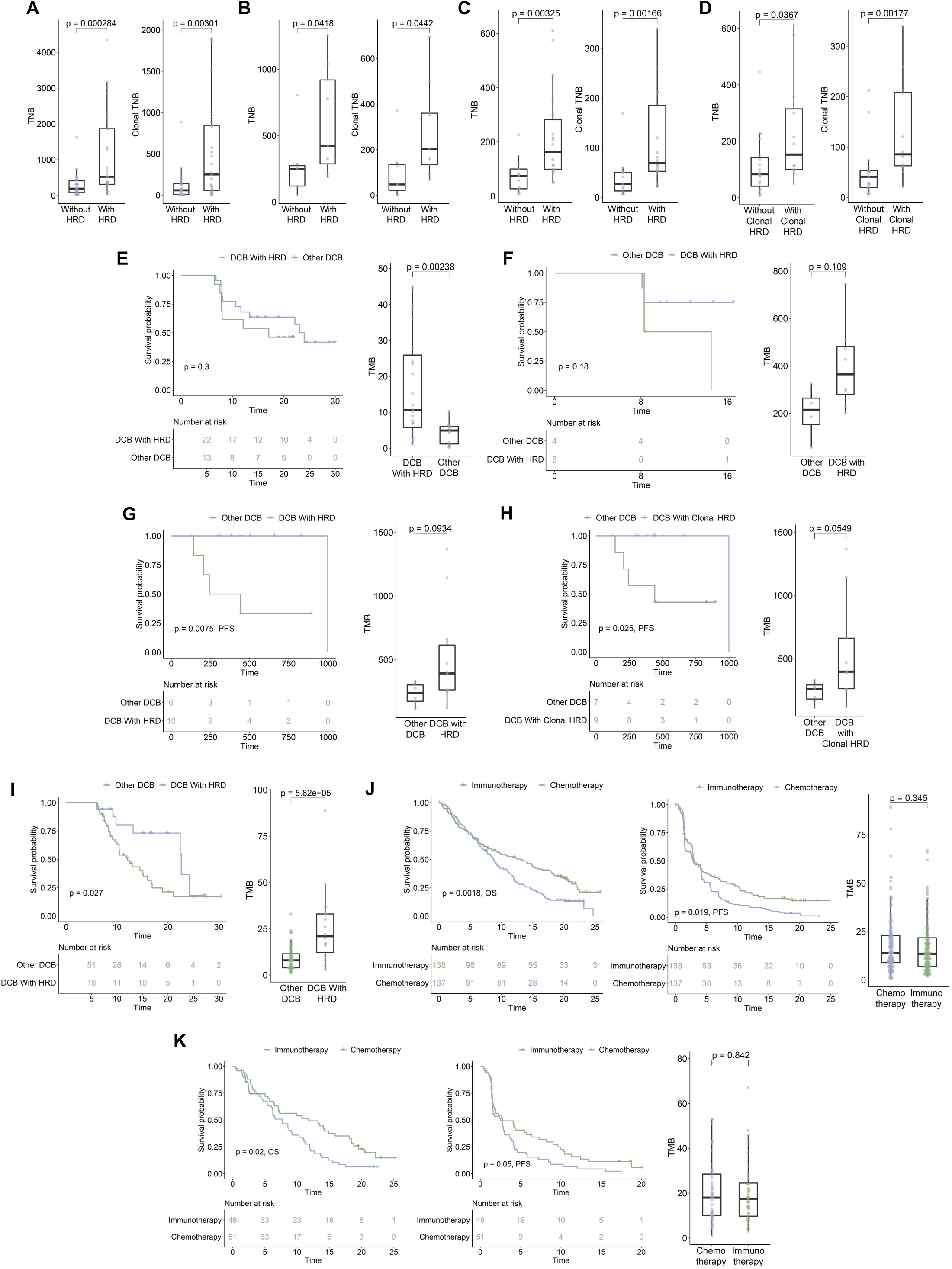
Integrated analyses on TNB, TMB and survival information of stratified patients from public cohorts. (A) Global and clonal TNB value distribution between non-squamous patients stratified by HR pathway mutations in Cancer Cell. 2018 dataset. Left: TNB. Right: clonal TNB. (B) Similar with (A), but on squamous patients. (C) Similar with (A), but in all patients from Science. 2015 dataset. (D) Similar with (C), but using HR pathway gene clonal mutations as the classification strategy. (E) PFS and TMB of all DCB patients with and without HR gene mutations in Cancer Cell. 2018 cohort. (F) Similar with (E), but in adenocarcinoma patients from Science. 2015 cohort. (G) Similar with (E), but in patients from Nat Genet. 2018 cohort. (H) Similar with (G), but using HR pathway gene clonal mutations as the classification strategy. (I) Similar with (G), but in patients from J Clin Oncol. 2018 cohort. (J) Survival and TMB of all HR pathway-mutant patients treated with immunotherapy and chemotherapy. Left: OS. Middle: PFS. Right: TMB distribution. (K) Similar with (J), but in patients with HR gene clonal mutations.

HR gene mutational information was further incorporated in classification to study the survival disparateness on public cohorts. For the adenocarcinoma patients from Science. 2015 dataset, subgroups with HR gene global and clonal mutations exhibited better progression-free survival (PFS) (Suppl. Fig. 8F). Similar observations could be made on PFS (Suppl. Fig. 8G) and overall survival (OS) (Suppl. Fig. 8H) data in Nat Genet. 2018 cohort. As for the targeted-sequencing cohort J Clin Oncol. 2018, patients possessing HR gene mutations consonantly got better PFS (Suppl. Fig. 8I) but incongruence on survival existed between adenocarcinoma (Suppl. Fig. 8J) and squamous (Suppl. Fig. 8K) patients. Again, when applying HR gene mutation as immunological biomarker in targeted sequencing, the sequencing panel should be elaborately designed. Additionally, DCB patients possessing HR pathway gene mutations surprisingly evinced better PFS and higher TMB than other DCB patients in Cancer Cell. 2018 (Fig. 6E) and Science. 2015 (Fig. 6F) datasets. Such group-wise discrepancy on PFS and TMB expanded in Nat Genet. 2018 dataset (Fig. 6G) and was analogously observed in OS data (Suppl. Fig. 8L). When narrowing to clonal HR gene mutations, the trend preserved in PFS (Fig. 6H) and OS (Suppl. Fig. 8M). Similarly, DCB patients with HRD manifested significantly better survival and higher TMB values than other DCB participants in J Clin Oncol. 2018 (Fig. 6I) dataset. Our multi-cohort analyses confirmed that HRD patients achieved generally better survival and the potency of HRD event in survival stratification, even among better responders in immunotherapy, i.e. DCB NSCLC patients.

With advantages including non-invasive, relatively costless, riskless and biopsy bias-reducing, liquid biopsies have received growing attention in tumor early diagnosis, recurrence monitoring and therapeutic guidance. We next conducted survival analysis in the publicly available blood-based Nat Med. 2018 dataset measuring circulating free DNA (cfDNA) to shed light on applying HRD mutational testing in blood biopsy. Firstly, no survival but significant TMB difference was observed between HR mutant and HR wild type chemo-treated patients in non-squamous (Suppl. Fig. 9A) and squamous (Suppl. Fig. 9B) patients, which denoted the uselessness of HRD as therapeutic biomarker in NSCLC chemotherapy treatment. When focusing on patients treated by immunotherapy, HR gene mutant group significantly got higher TMB but inconspicuous group-wise survival difference existed in non-squamous (Suppl. Fig. 9C) and squamous (Suppl. Fig. 9D) subpopulation. Regarding these HRD events were measured by targeted sequencing, different results could possibly be observed by more comprehensive HRD testing strategy. Ultimately, for patients with HR gene global and clonal mutations, patients received immunotherapy exhibited significantly better survival (Fig. 6J-K). Apart from prioritizing patients with better immunological response, HRD event could provide suggestions over different therapy types for NSCLC patients.

Last of all, we investigated the mutational frequency of HR genes in multiracial datasets to guarantee the universality of HRD event in untreated NSCLC. The HR and core HR pathway gene global and clonal (defined by Taf>=0.3) mutation frequency in Sci Rep. 2015 Chinese SCC (Suppl. Fig. 10A), J Thorac Oncol. 2020 African American NSCLC (Suppl. Fig. 10B), TCGA-LUAD (Suppl. Fig. 10C) and TCGA-LUSC (Suppl. Fig. 10D) cohorts were calculated. In addition, HR pathway gene alternation information including mutations, deep deletions and epigenetic silencing from a previous paper^14^ were retrieved for TCGA-LUAD (Suppl. Fig. 10E) and TCGA-LUSC (Suppl. Fig. 10F) datasets. Both histological subtypes exhibited around 50% HR gene alternation frequency. The quantified HRD event (HRDscore) additionally demonstrated congruent strong positive correlation with CNV burden in two TCGA lung cancer datasets (Suppl. Fig. 10G-H), again validated the concomitant relationship between HRD event and drastic CNV. Together with discoveries listed above, HRD events occurred more frequently in better-responded patients receiving immunotherapy regardless of the treatment regimen and the clinicopathologic characteristics. We observed the resultant higher TMB, TNB and consequently longer survival brought by HRD as well as the substantial amount of HRD events in multiracial samples, which all countenanced the potential of HRD testing as a novel immunotherapy biomarker.

## Discussion

Lung cancer is the major source of worldwide cancer-related deaths. Superior than surgery and adjuvant chemotherapy that failed to completely eliminate the recurrent risk^36^, the emerging neoadjuvant therapy improves patients’ survival. In this study, 3 adenocarcinoma and 11 squamous cell carcinoma NSCLC patients were enrolled to receive immuno-neoadjuvant therapy with no obvious duration disparity and the inner genetic cause of response difference was further characterized by WES. A set of genes were altered and HR pathway gene clonal deactivation gave rise to prevalent HRD events in MPR group, which orchestrated TMB, mutational as well as SCNA level ITH and eventually contributed to higher clonal TNB generation. Validations in public cohorts containing NSCLC patients with different treatment design and clinicopathologic characteristics further confirmed the aggregation of HR pathway gene alternations in better-responded population. Higher TMB, TNB and better survival in HR-mutated patients was additionally observed and statistics on the HR pathway gene alternation frequency in multiethnic cohorts eventually guaranteed the universality of HRD testing in immunotherapy patient stratification. To our knowledge, our work is the first report to state the strong association between neoadjuvant immunotherapy benefit and HRD event in NSCLC. We also provided the first comprehensive investigation on the association between HRD and lung cancer therapies. The inspection of HR pathway gene status could serve as novel or complementary indicator directing immuno-neoadjuvant and immunotherapy treatment decisions for NSCLC patients.

Through comprehensively analyses on genetic alternations, biological characteristics of the patients with distinct therapeutic responses were further elucidated. Numbers of driver genes were found mutated in patient groups and mutations of EGFR/ALK/LKB1 was absent in all samples. For mutational comparisons with other cohorts, cancer driver genes including *ZFHX3*, *PIKACA* and *NUP93* exhibited deviating mutational frequency. *ZFHX3* mutational status was reported as a protective marker for NSCLC immunotherapy^37^ recently while *PIK3CA* mutation therapeutically benefits microsatellite stable colorectal cancer patients^38^. Though *PIK3CA* mutations activating PI3K/AKT/mTOR pathway resulted in T cell-mediated immunotherapy resistance^39^, our observation highlighted the bi-nature of *PIK3CA* SNVs. Besides, group-specific altered cytobands were prioritized through comparisons and both 9p13.3 and 12q24.31 demonstrated inverse alternation trend in MPR/Non-MPR patients. Patients with amplified 9p13.3 tend to have worse progression-free survival in prostate cancer^40^ while 12q24.31 gain was reported to be a powerful marker in neuroblastoma progression^41^. The alternation trends of 9p13.3 and 12q24.31 in our samples shared high congruity with these known mechanisms. Additionally, genes including *MLF1*, *TIPARP* and *WWTR1* showed high focal SCNA group-specificity. Both *MLF1* and *TIPARP* were inextricably linked to tumor suppression mechanisms^42, 43^, matching the fact that MPR patients better responses to immuno-neoadjuvant therapy. Finally, by intersecting the group-specific cytobands from GISTIC and genes possessing focal SCNA, six genes including *CASC5* and *RAD51* were pinpointed. *CASC5* was previously identified as tumor driver in LUAD^44^ and the elevation of RAD51 showed worse survival in both LUAD and SCC^45^. Our analyses demonstrated the association between their SCNA and better neoadjuvant immunotherapy response. All in all, multiple group-specific alternations were identified, which shed light on the mechanisms beyond therapeutic response and presented potential biomarkers with indicative ability in therapy outcome prediction.

The HRD events concomitantly causing augmented genomic alternations are frequently observed in breast and ovarian cancer but study reporting their existence and role in lung cancer is rare. Though two recent work reported HRD prevalence in selected NSCLC patients^46^ and the association between HR gene alternations and elevated TMB as well as prolonged PFS in patients treated with immune checkpoint inhibitors (ICIs)^47^, our work, for the first time, proposed that the NSCLC neoadjuvant therapy benefit is associated with HRD status and the generated TMB, ITH and clonal TNB alternations, regardless of the treatment regimen. Instead of an observation on specific study, our comprehensive analyses incorporating plentiful public data originally validated the consistency of the association between HRD event and lung cancer therapy outcomes. Though germline *BRCA* gene mutation testing is a well-established HRD event biomarker^48^ and has been widely applied to assess breast and ovarian cancer risks for the use of platinum-based chemotherapy, poly ADP-Ribose polymerase (PARP) inhibitors and DNA damaging reagents, our results concluded the HRD events were driven by somatic HR pathway gene alternations in NSCLC and emphasized the importance of somatic HR pathway genetic testing in NSCLC immuno-neoadjuvant and immunotherapy treatment determination. We further accentuated the importance of sequencing panel design when applying HR pathway gene diagnostic testing as biomarker, providing innovational direction for future medical transformation.

PD-L1 expression, being the only FDA-approved biomarker for LUAD patients receiving ICIs^49^, could become unwarranted in ICI response prediction due to the influence of molecular as well as clinical characteristics on PD-L1 state. In lung cancer neoadjuvant immunotherapy, sporadic cases reported the association between high PD-L1 expression and pCR state^50^ and a recent study discovered the possible correlation between PD-L1 and MPR as well as pCR rate in meta-analysis^51^. As for our discoveries, the PD-L1 expression failed to prioritize MPR patients. Concerning the variability of PD-L1 expression conferred by molecular attributes, we speculate the testing on HRD event or HR pathway gene alternations could aid the MPR patient stratification in NSCLC neoadjuvant immunotherapy, especially for the PD-L1 negative subpopulation.

As mentioned above, we proposed the hypothesis that the provoked HRD events eventually compensate clonal neoantigen generation through SCNA. Previous reports linked neoantigen generation to the genome instability caused by various genetic alternations including germline mutations on mismatch repair (MMR), *POLE* and *BRCA* genes^52^ in mouse models. For example, the resultant continuous neoantigen generation in colorectal and pancreatic mouse cancer cells with DNA MMR was observed by Germano et al^53^. However, scarce research focused on the HRD event and the association between neoantigen generation, especially in lung cancer. Our work originatively proposed the novel association between SNV/SCNA ITH provoked by HRD event and the neoantigen elevation in lung cancer, yielding implications for tumor precision medicine.

However, some issues should be noted. Albeit key analytical metrics reached significant, all the conclusions on neoadjuvant therapy were derived from a relatively small patient amount, an increased size of patient enrollment could better generalize our conclusions. Besides, instead of OS requiring a much longer follow-up, we utilized MPR, which is significantly predictive of long-term OS in neoadjuvant therapy^54^, as a surrogate endpoint for therapy response evaluation. Associations between HR gene deficiency and longer survival was unveiled in non-neoadjuvant immuno/chemotherapy studies^46, 47^, our study highlighted the prophetic power of HRD events in neoadjuvant immunotherapy. As a final point, main conclusions in this study were derived from WES data on FFPE/fresh tissues. Further incorporation on matched blood samples in sequencing could solidate our discoveries and better surveil the HRD event in blood, hence aid future translational medicine application.

## Conclusions

Our study associated the deactivation of HR pathway genes and the resulting HRD event with the improved therapeutic outcomes of immuno-neoadjuvant treatment in lung cancer. HRD events orchestrated TMB as well as multi-aspect ITH and compensated clonal neoantigen generation in our samples. Discoveries were further validated by public cohorts. In summary, the inspection of HR gene genetic status could serve as unprecedented guide prior to the armamentarium of immuno-neoadjuvant and immunotherapy treatments for NSCLC patients.

## Data Availability

Data generated in the current study is available from the corresponding author on reasonable request. Data used in the validation analyses were either downloaded from the supplemental files of the publications or from cBioPortal database.

## List of abbreviations

CNV: copy number variation
CR: complete response
DCB: durable clinical benefit
DDR: DNA damage repair
HR: homology-dependent recombination
HRD: homologous recombination deficiency
InDel: small insertions and deletion
ITH: intratumor heterogeneity
LUAD: lung adenocarcinoma
LUSC: lung squamous cell carcinoma
MPR: major pathological response
NDB: no durable benefit
NSCLC: non-small-cell lung cancer
OS: overall survival
pCR: pathological complete response
PFS: progression-free survival
PR: partial response
SCC: squamous cell carcinoma
SCNA: somatic copy number alteration
SNV: somatic single nucleotide variant
TMB: tumor mutational burden
TNB: tumor neoantigen burden
WES: whole-exome sequencing

---

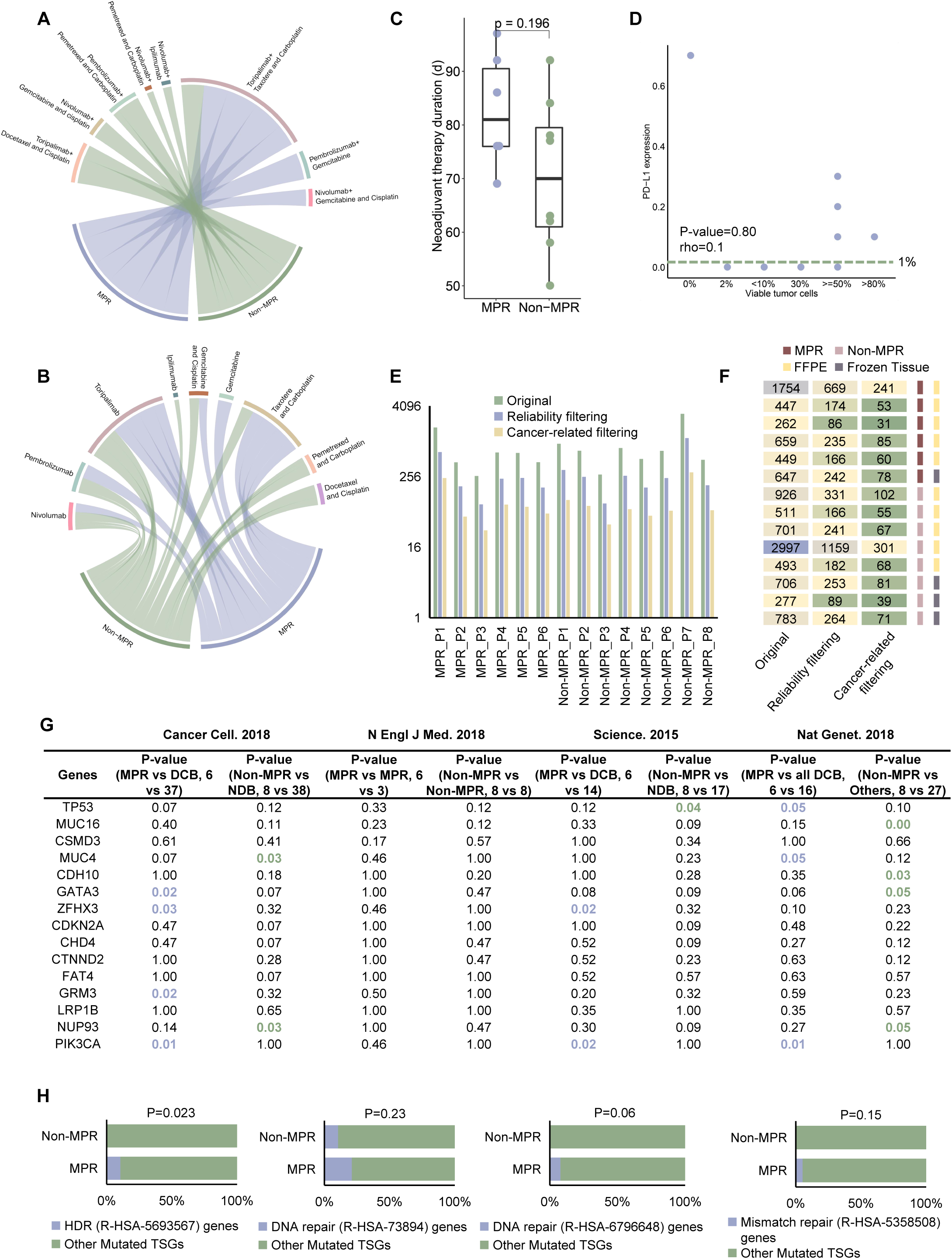

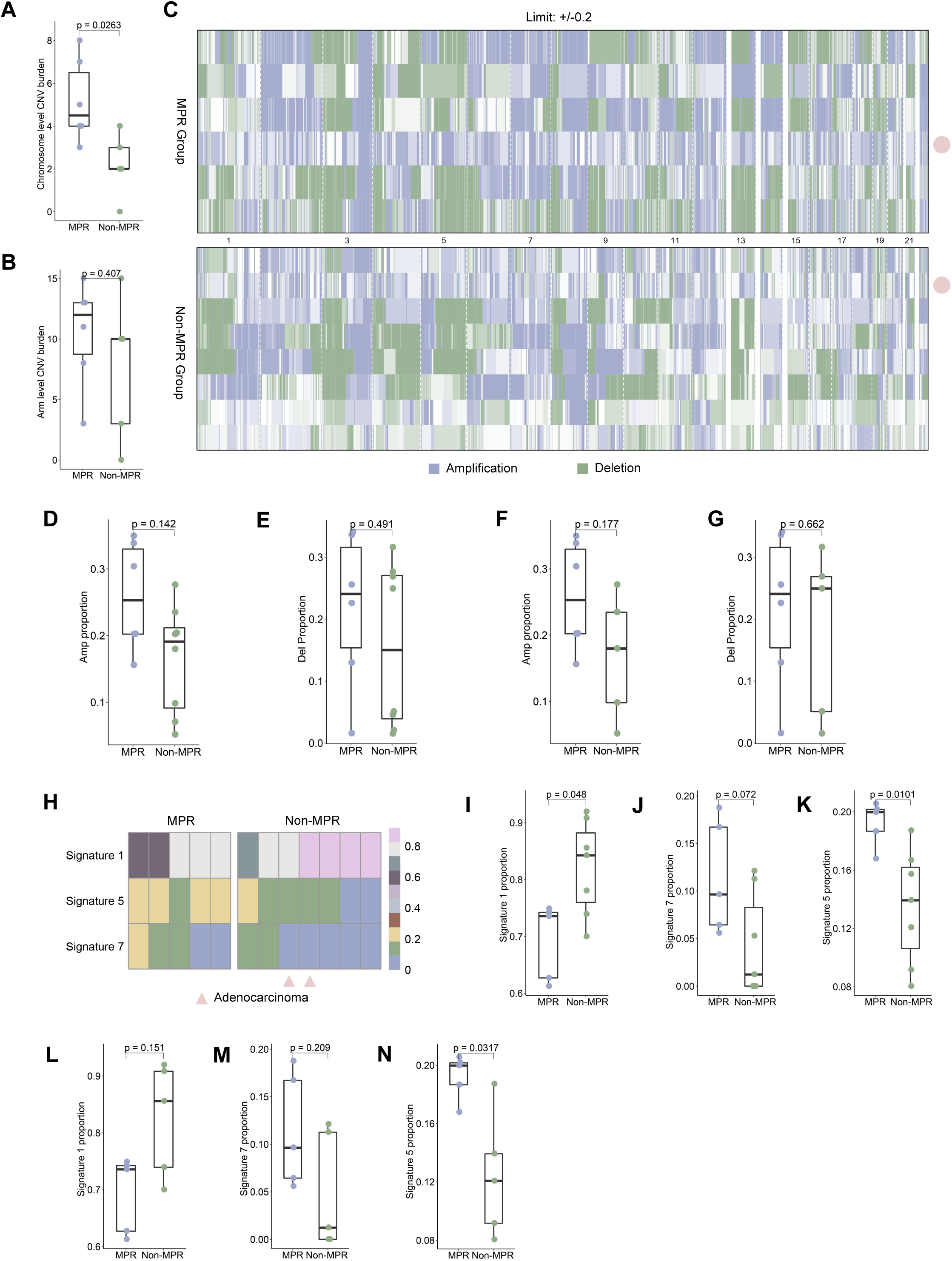

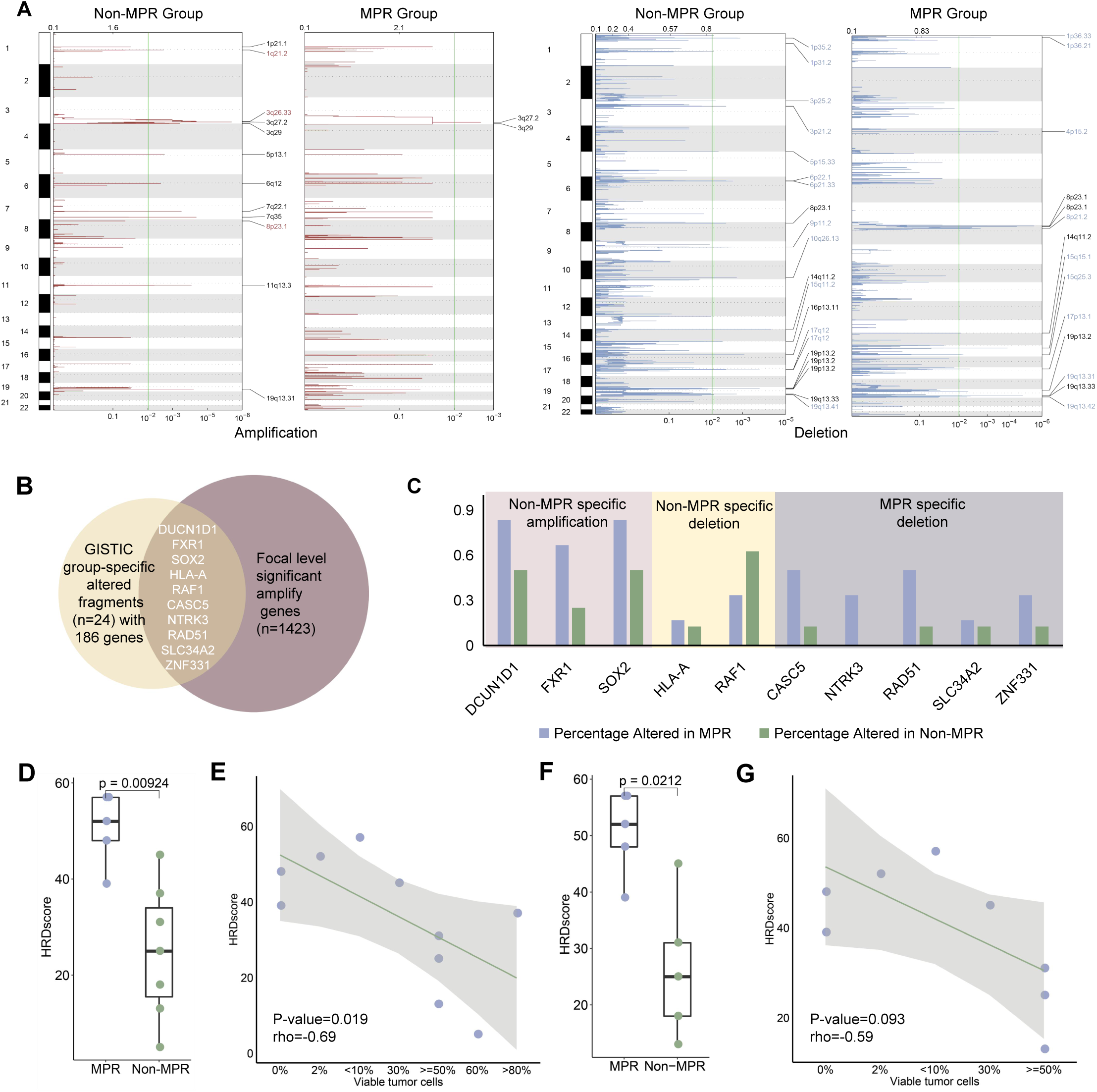

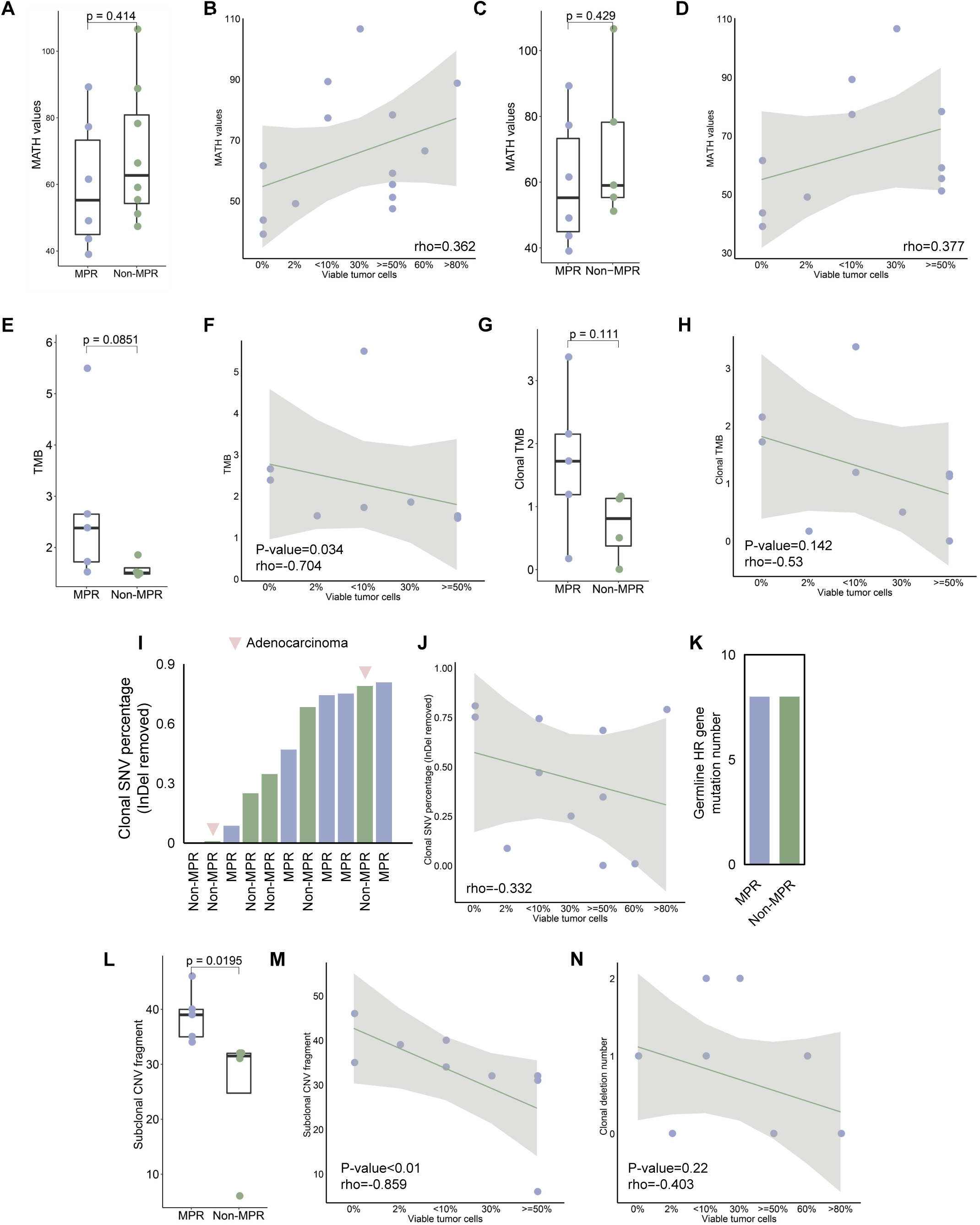

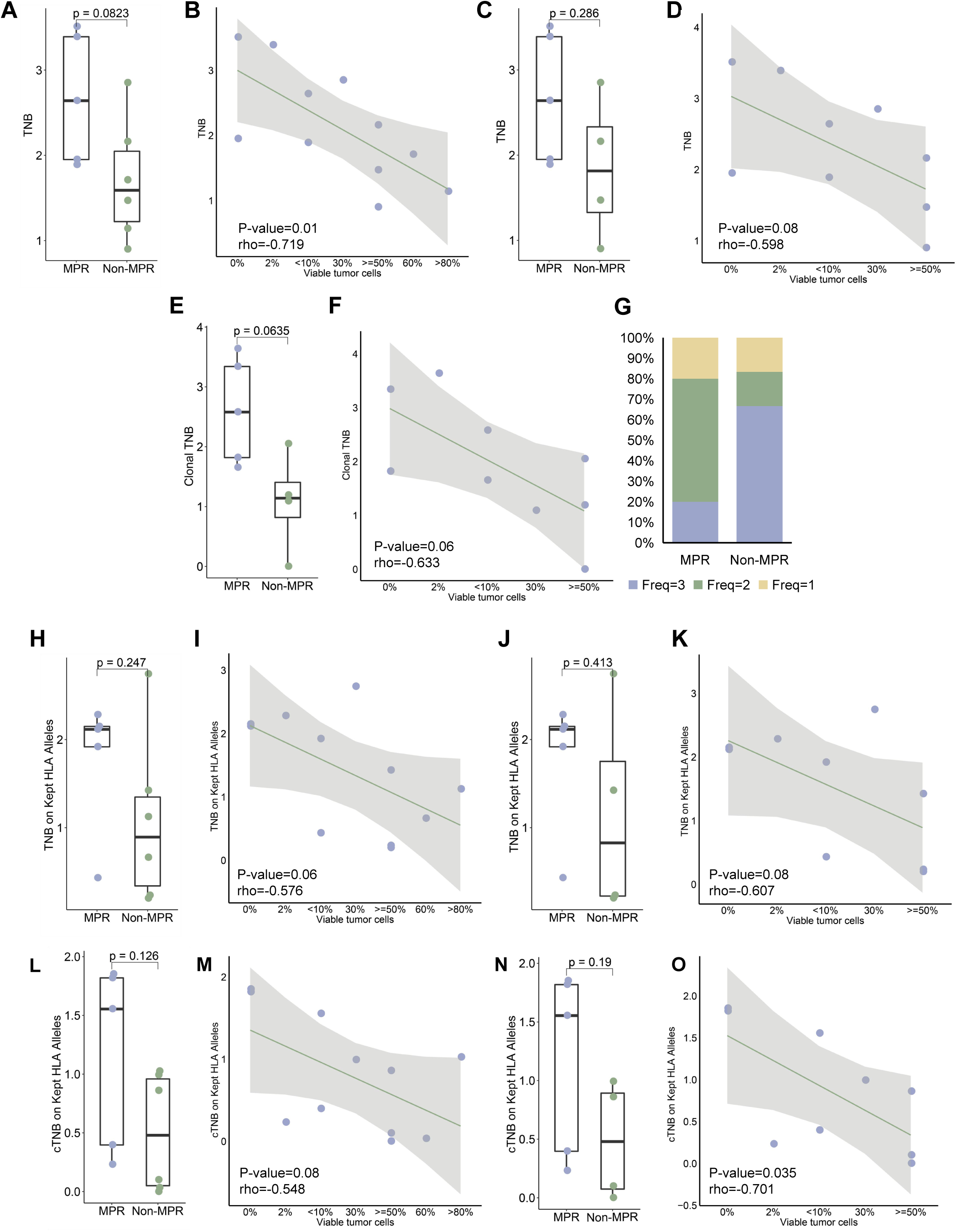

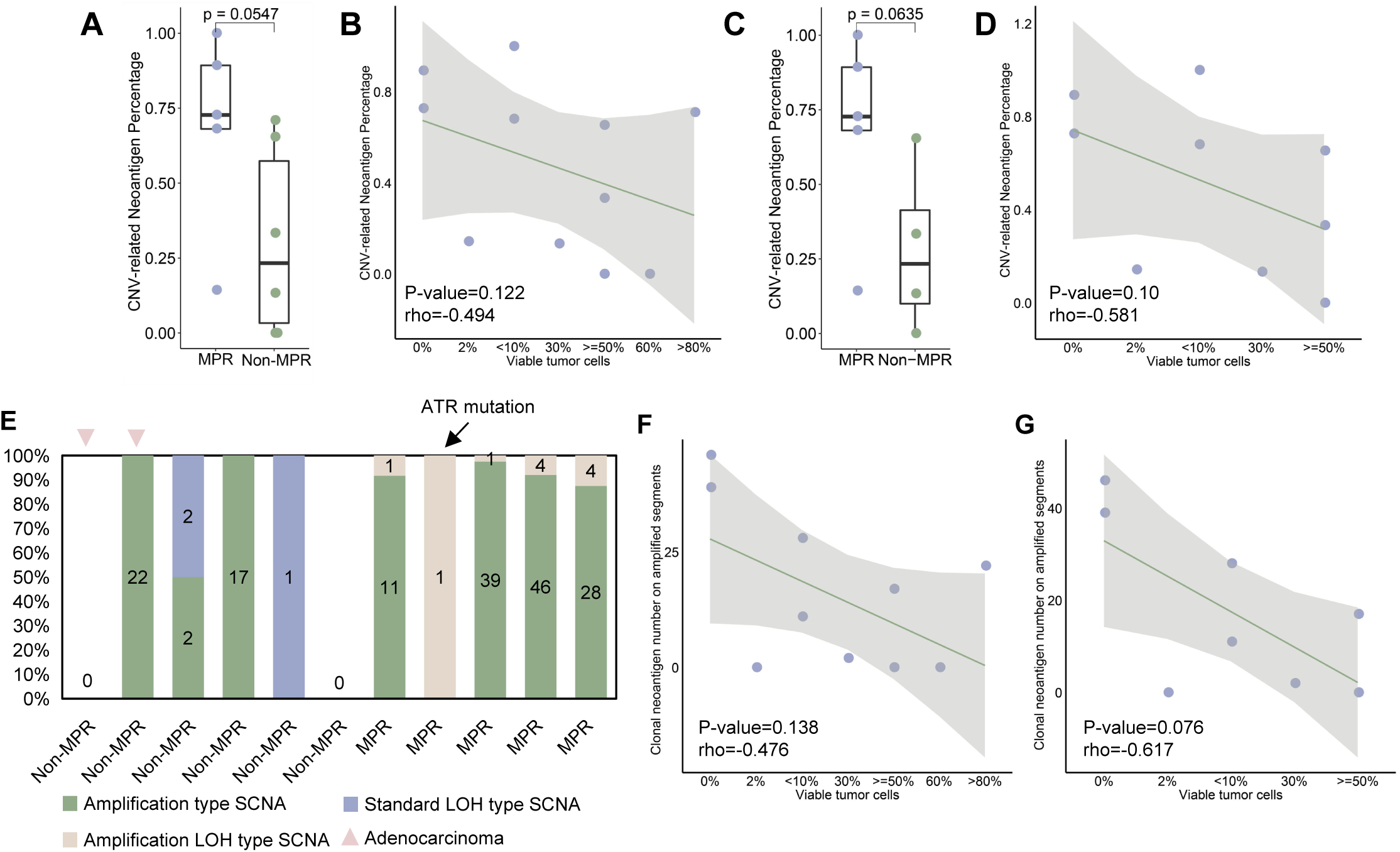

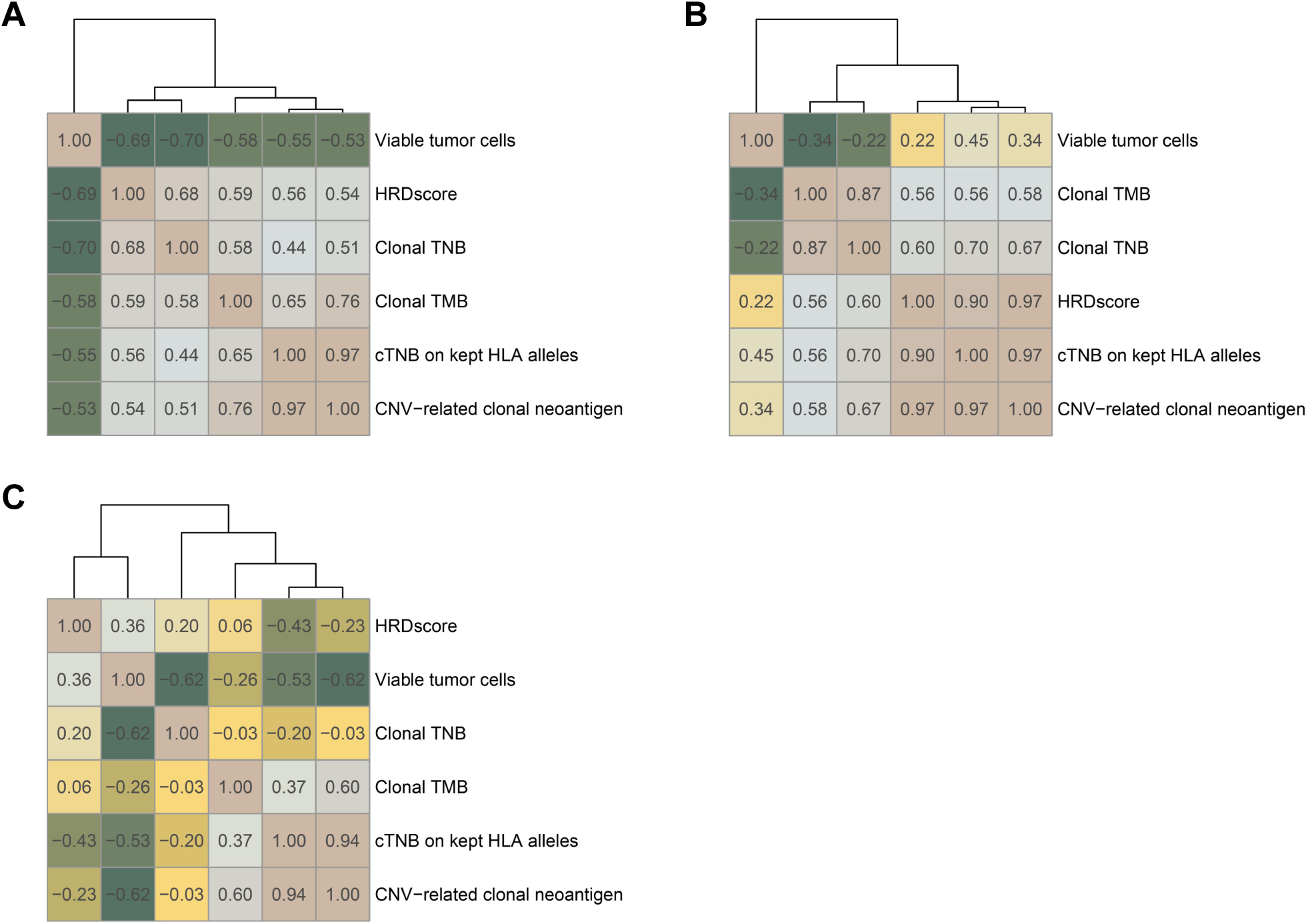

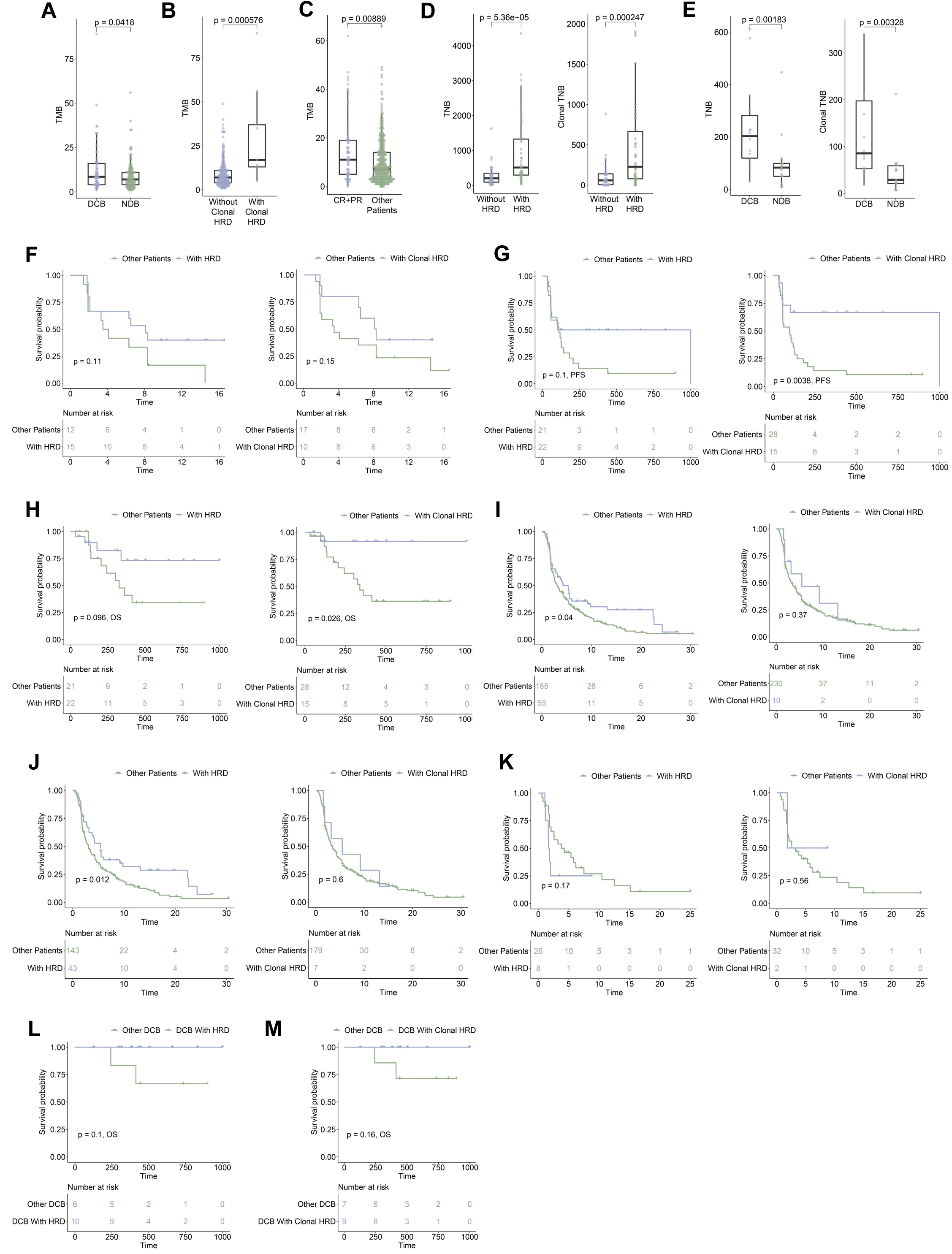

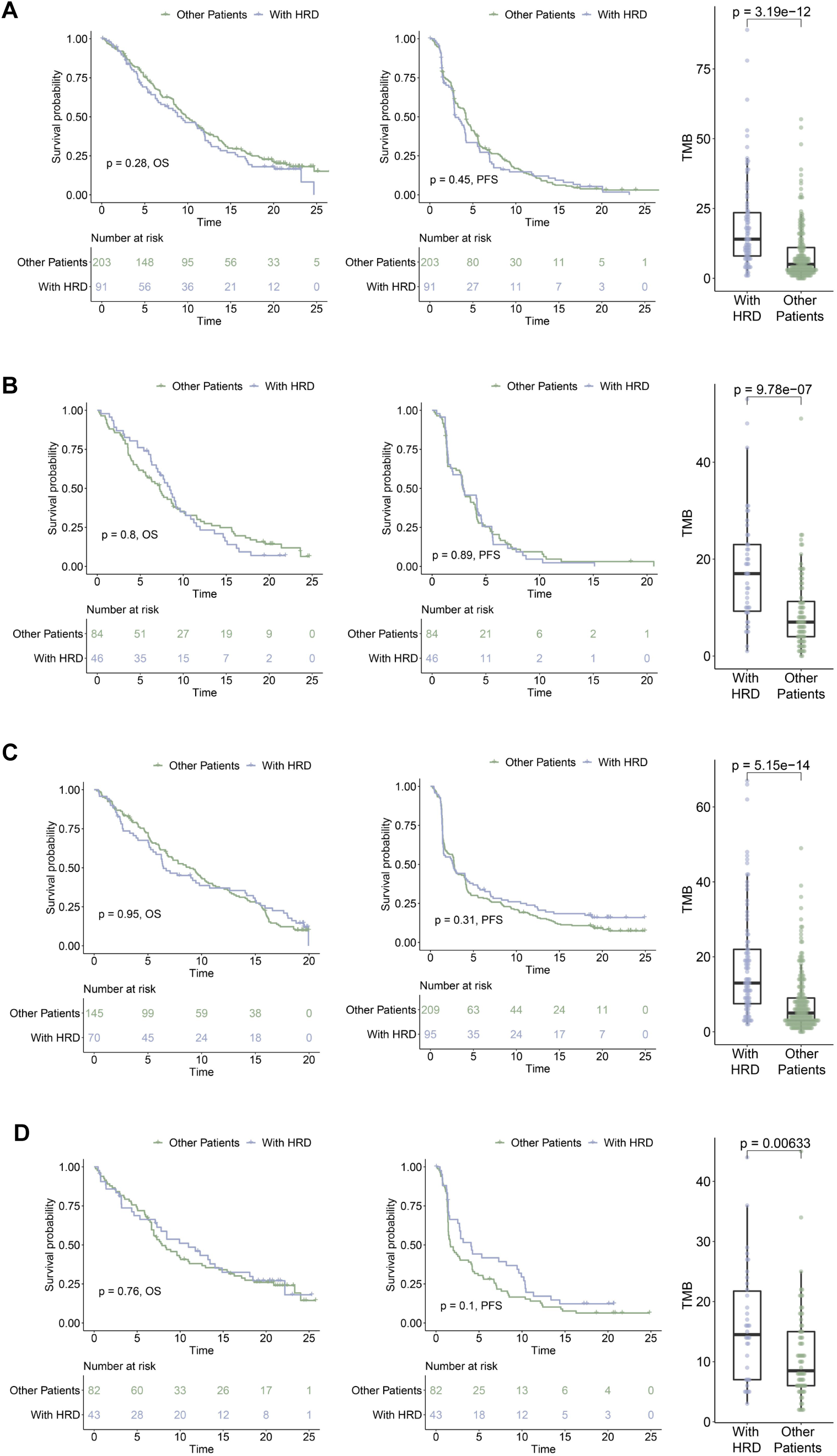

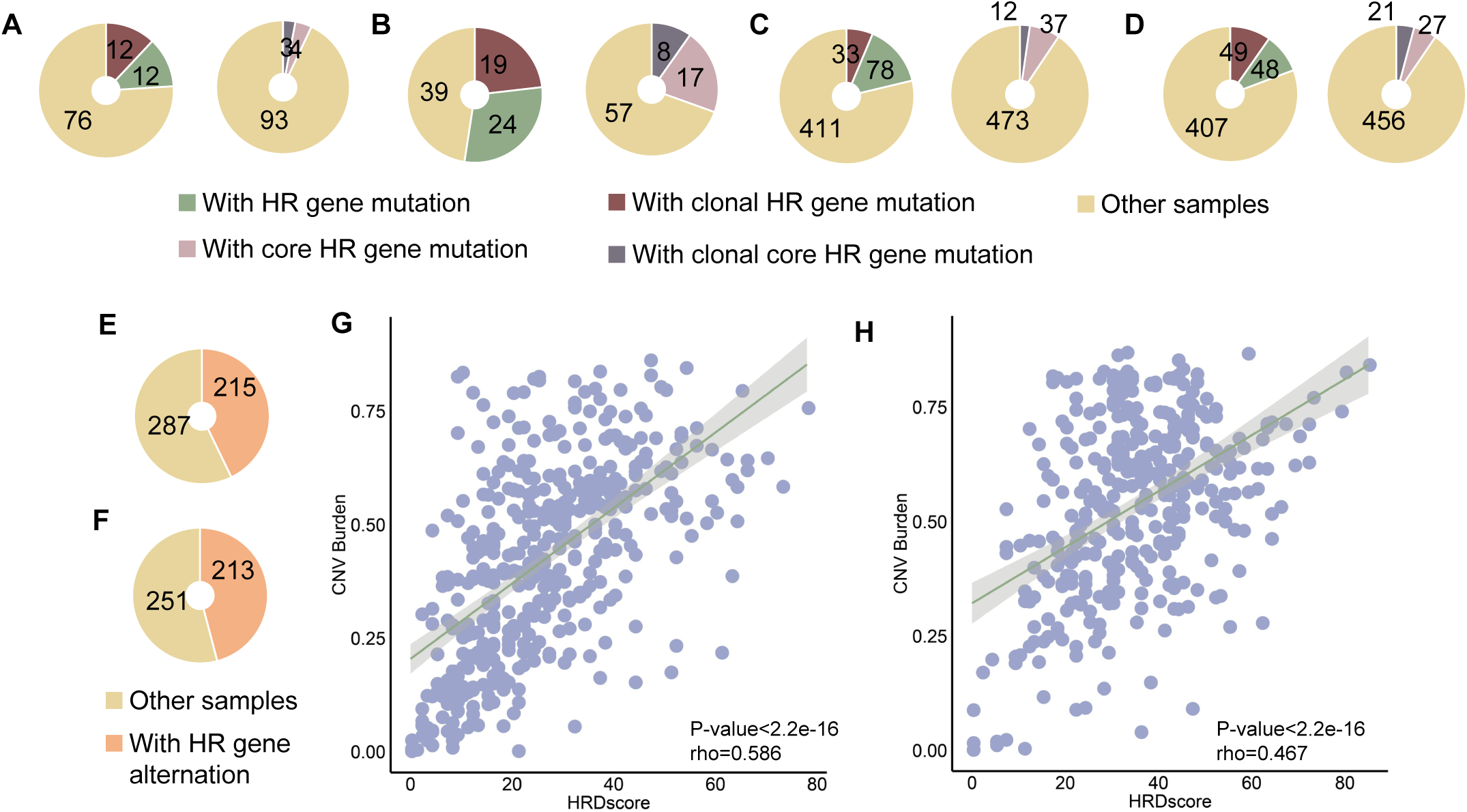

